# Longitudinal study of gesture decoding in a clinical trial participant with ALS

**DOI:** 10.1101/2025.09.26.25335804

**Authors:** Daniel N. Candrea, Miguel Angrick, Shiyu Luo, Rohit Ganji, Christopher Coogan, Griffin W. Milsap, Kathryn R. Rosenblatt, Alpa Uchil, Lora Clawson, Nicholas J. Maragakis, Mariska J. Vansteensel, Francesco V. Tenore, Nicholas F. Ramsey, Matthew S. Fifer, Nathan E. Crone

## Abstract

Brain-computer interfaces (BCIs) have the potential to preserve or restore communication and device control in people with paralysis from a variety of causes. For people living with amyotrophic lateral sclerosis (ALS), however, the progressive loss of cortical motor neurons could theoretically pose a challenge to the stability of BCI performance. Here we tested the stability of gesture decoding with a chronic electrocorticographic (ECoG) BCI in a man living with ALS and participating in a clinical trial (ClinicalTrials.gov, NCT03567213). We evaluated offline decoding performance of attempted gestures over two periods: a 5-week period beginning roughly 2 years post-implant and a 6-week period ending roughly 5 months later. Decoder sensitivity was high in both periods (90 - 98%), while classification accuracy was 37 - 68% in the first period and worsened to 23 - 39% in the second. We investigated multiple frequency bands that were used as model features in both periods, and we observed reductions in high gamma band power (70 - 110 Hz) and between-class separation during the second period compared to the first. Over the 5-month period motor function did not appreciably decline. These results, albeit preliminary, suggest that declines in the neural population responses that drive ECoG BCI performance can occur without overt signs of disease progression in people living with ALS, and could serve as a biomarker for disease progression in the future.

## Introduction

Amyotrophic lateral sclerosis (ALS) is a neurodegenerative disease of the upper and lower motor neurons that can result in locked-in syndrome (LIS), a condition in which muscle weakness limits mobility and communication in spite of normal cognition^1^. Because eye movements are usually spared at first^2,3^, eye-trackers are commonly used to control assistive devices, but may cause fatigue^4–6^ and become unreliable as eye movements eventually deteriorate^3,7–9^.

Alternatively, people living with ALS can potentially control assistive devices using brain computer interfaces (BCIs), which detect and translate the neural correlates of attempted actions into device-specific outputs^10–15^. Recent studies have demonstrated that ECoG BCIs can offer long-term stability without frequent model retraining^10,15,16^ or recalibration^13^. This long-term stability may be due to the summation of responses from the large populations of neurons captured by EcoG recordings^17–19^. For example, we have shown that electrocorticographic (ECoG) modulations during attempted grasping movements can be used to make “brain-clicks” to control a switch-scanning spelling application over a period of months^15^ and years^10,20^.

On the other hand, multiclass gesture decoding for people with ALS has been explored only recently, despite potential BCI users scoring it favorably as a possible control strategy^21^. For example, classification of upper limb, lower limb, and/or facial gestures in participants with ALS has been demonstrated using micro-electrode arrays (MEAs)^22^ and endovascular stent-electrode arrays (Stentrodes)^11^, the latter of which primarily target lower limb cortical representations.

As part of our CortiCom clinical trial (ClinicalTrials.gov Identifier: NCT03567213), we investigated the feasibility of decoding gestures over multiple months from a clinical trial participant with ALS that was implanted with an ECoG-based BCI. We collected neural activity from 16 attempted upper limb, lower limb, and facial gestures over two periods: a 5-week period beginning roughly 2 years post-implant and a 6-week period ending roughly 5 months later. We also developed an algorithm for selecting an optimal subset of six “control gestures” that could simulate an online BCI control strategy. These “control gestures” were selected based on their performance in a model training and validation phase and were then tested offline using held-out testing data.

We observed a decline in offline decoding performance over the course of 5 months. Upon comparing the neural characteristics of attempted gestures from both periods, we observed a corresponding decrease in the magnitude of task-related spectral responses in multiple frequency bands, including the high gamma band (70 - 170 Hz in this study). Over the same period, monthly neurological examinations did not demonstrate a significant decline in motor function. A prior longitudinal study showed that signal strength and BCI performance can decline in patients with LIS due to advanced ALS, in whom there is no measurable strength in the upper limbs^20^. In contrast our participant had residual, albeit impaired, strength in his upper limbs, allowing serial clinical measurements. Our study thus provides important evidence that ECoG BCI performance and signal strength can decline in a person living with ALS in the absence of an observable change in their clinical exam. Moreover, our findings suggest that ECoG BCI performance and signal strength could potentially serve as a more sensitive biomarker for disease progression than clinical examination alone.

## Methods

### Clinical trial

This study was conducted as part of the CortiCom clinical trial (ClinicalTrials.gov Identifier: NCT03567213). Results related to the primary outcome variables were reported in our previous work and were related to the safety and recording viability of the implanted device, as well as the BCI functionality enabled by the implanted device^13,15^. In this study, we do not report results related to the primary outcome variables. The study protocol was reviewed and approved by the Johns Hopkins University Institutional Review Board (IRB) and by the US Food and Drug Administration (FDA) under an investigational device exemption (IDE). Clinical trial inclusion and exclusion criteria are available in Supplementary Method 1.

### Participant

All results reported in this study were based on data from the first participant (CC01) in the CortiCom trial. The participant was informed of the nature of the research and implant related risks and gave written consent to participation in the trial. The participant was a right-handed man in his 60s at the time of enrollment. He was diagnosed with ALS 8 years prior and had progressive dyspnea as well as progressive dysarthria and swallowing impairments due to bulbar dysfunction. The participant did not rely on assistive communication devices because he could still produce overt speech, though very slowly and with limited intelligibility, as reported previously^15^. Due to weakness in his upper limbs (Medical Research Council (MRC) scores ranging from 2 to 4), he was incapable of performing any activities of daily living without assistance. He had insufficient strength to hold a cup with one hand but could partially close his fingers when attempting to grasp. He could ambulate due to good strength in his lower limbs, but he had significant gait imbalance with risk of falls due to impaired arm swing, requiring supervision to prevent fall-related injuries. The ALSFRS-R score was used to periodically evaluate the participant’s functional disability. Regarding this study, an assessment was taken one week before the first multi-week gesture recording period (Supplementary Note 1).

### Clinical strength measurements

Throughout the CortiCom clinical trial the participant underwent a full neurological examination by the same board-certified neurologist approximately once a month. Each examination included clinical strength measurements for 14 movements: neck flexion, neck extension, shoulder abduction, elbow flexion, elbow extension, wrist flexion, wrist extension, finger abduction, hip flexion, hip extension, knee extension, knee flexion, foot dorsiflexion, and foot plantar flexion. For each movement, strength was rated on the MRC scale: 5 - normal resistance, 4 - reduced resistance, 3 - ability to move only against gravity, 2 - inability to resist gravity, can move with gravity negated, 1 - minimal movement (twitch), and 0 - no observable movements. All lower extremity movements were rated 5/5 throughout the study, without any observable deviation from baseline. MRC ratings for movements in the neck ranged from 4 to 5 and ratings for upper extremities ranged from 2 to 4 and did not exhibit any consistent decline throughout the study (Supplementary Fig. 1).

### Neural implant

The CortiCom study device consisted of two 8×8 subdural high-density (HD) ECoG grids (PMT Corporation; Chanhassen, MN) connected to a percutaneous 128-channel Neuroport pedestal (Blackrock Neurotech Corporation; Salt Lake City, UT). Final assembly and sterilization of the study device was performed by Blackrock Neurotech. Each grid consisted of platinum-iridium disc electrodes (0.76 mm thickness, 2 mm diameter exposed surface) with 4 mm center-to-center spacing embedded within a soft silastic sheet with a total surface area of 12.11 cm^2^ (36.6 mm x 33.1 mm). The ECoG grids were implanted subdurally over the left hemisphere, over sensorimotor cortex representations for speech and upper extremity movements (Supplementary Fig. 2). The device included two subdural reference wires, each with an uninsulated tip that matched the surface area of a recording electrodes, placed on top of the ECoG grids. Signals were always referenced to the same reference wire. During all recordings, a small (24.9 mm x 17.7 mm x 17.9 mm) detachable head stage (Neuroplex-E, Blackrock Neurotech Corporation) was connected to the percutaneous pedestal for signal amplification, digitization, and digital transmission via a micro-HDMI cable to the Neuroport Biopotential System (Blackrock Neurotech Corporation).

### Data collection

Data was collected during 20 sessions (study visits) between 747 and 897 days after device implantation (149 days; ~5 months). More specifically, data from sessions 1 - 10 were recorded between days 747 and 784 (24 - 25 months post-implantation) and data from sessions 11 - 20 were recorded between days 855 and 897 (28 - 29 months post-implantation). We denote these periods as Periods 1 and 2 and denote the data collected in these sessions as *D*^1^ and *D*^2^, respectively.

Five recording blocks were collected during each study visit, where each block was ~8 min in duration. For each recording block, the participant was instructed to attempt brief gestures in response to visual cues that were presented as text on a computer screen (Supplementary Fig. 3a). The gestures included 12 attempted movements of the right upper limb (contralateral to the implanted grids), 3 attempted facial movements, and 1 attempted movement of the right foot. The participant was instructed on the gesture to perform for each text cue. Due to the participant’s weakness, the gestures were performed with varying degrees of impairment (Supplementary Table 1).

Neural signals were recorded by the Neuroport system at a sampling rate of 2 kHz after an anti-aliasing low pass filter of 500 Hz was applied. BCI2000 was used for data collection^23^.

In each block, each of the 16 gestures was repeated four times (in total, 4 trials/gesture x 16 gestures = 64 trials/block) in a pseudo-random interleaved fashion (Supplementary Fig. 3b). Each trial consisted of a visual cue, followed by an interstimulus interval (ISI), during which the participant remained still and fixated his gaze on a crosshair in the center of the monitor. We set the length of the visual cue to 2 s to allow the participant sufficient time to read which movement he was instructed to attempt. The ISI varied uniformly between a lower and upper bound (4 - 6 s) to reduce anticipatory behavior. In total, ~800 min of data (400 trials/gesture split evenly between Periods 1 and 2) were collected.

### Feature computation

#### Calibration

Similarly to previous work^15^, a 30 s calibration period was recorded during each study visit, from which the statistics of the spectral-temporal log power of each frequency bin were used for standardization of power estimates during model training and BCI operation.

#### Spectral power computation

We excluded channels from the grid covering the sensorimotor cortex representation of speech and facial movements from further analysis due to movement artifact observed in the data collected during Period 2. We therefore excluded attempted facial movements (Tongue, Grimace, and Eyebrows) from further analysis and analyzed the remaining 13 non-facial gestures. We also excluded channels 65, 73, 81, and 83 from the grid covering the upper limb sensorimotor cortex due to poor signal quality. (Supplementary Fig. 2). For each of the remaining 60 channels, signals were referenced to their common average. We used a Fast Fourier Transform (FFT) filter (Tukey window with a cosine fraction of 0.25) to compute the spectral power of 256 ms segments that were shifted by 100 ms increments. We computed the spectral power for the entirety of each block. For each frequency bin, the spectral power was log-transformed and standardized (z-scored) to the statistics (mean and standard deviation) of that frequency bin from the calibration period recorded on the same day. Due to standardization, the resulting log-power values (z-scores) were unitless.

#### Frequency band selection

We investigated a variety of spectral features for potential use in model training^24^. These included δ (delta): 0.5 - 4 Hz; θ (theta): 4 - 8 Hz; μ (Mu): 8 - 13 Hz; β_1_ (beta 1): 13 - 19 Hz; β_2_ (beta 2): 19 - 30 Hz; γ_low_ (low gamma): 30 - 50 Hz; γ_high1_ (high gamma 1): 70 - 110 Hz; γ_high2_ (high gamma 2): 130 - 170 Hz. For each frequency band, we summed the log-standardized spectral power between the corresponding low and high frequency bounds for each channel. We determined which frequency bands contained significant gesture-related modulation by computing the modulation at the peak (for γ_high1_, and γ_high2_ frequency bands, due to modulation in the positive direction) or nadir (for δ, θ, μ, β_1_, β_2_, and γ_low_ frequency bands, due to modulation in the negative direction) of the trial-averaged frequency band power for each channel. For any frequency band, if the peak or nadir z-scored trial-averaged modulation was greater or less than a z-score of 1.96 or −1.96, respectively (corresponding to an α significance value of 0.05), for any channel at any positive time point within the trial duration (−2 to 5 s relative to the onset of the visual gesture cue), this frequency band was deemed to contain significant gesture-related modulation. Otherwise, the frequency band did not contain significant gesture-related modulation and was excluded from further analysis. We performed this analysis using the data collected in Period 1 but not using the data collected in Period 2 because we had begun this analysis before Period 2. This means that for both Periods 1 and 2, we used the frequency band features identified in Period 1.

#### Feature generation

For each frequency band with significant modulation, a 60-channel feature vector was generated. The feature vectors from each frequency band were vertically concatenated to produce a *b*\*60-dimensional feature vector, where *b* was the number of frequency bands with significant gesture-related modulation. Once these frequency bands were determined, *b* remained constant throughout the entire analysis. The *b*\*60-dimensional feature vector was generated every 100 ms, due to the spectral time shift mentioned above. In the context of model training, validation, and testing, we will refer to these feature vectors as “samples” from now on.

### Training, validation, and testing

We trained multiple models with sets of training data from increasing numbers of sessions to 1) investigate the relationship between the amount of training data and model performance, and 2) account for possible variations in how gestures were attempted on a day-to-day basis. Day-to-day variations in gesture attempts could have been reflected in the neural signals (Supplementary Note 2). This led us to select an optimal subset of six gestures (control gestures) for each set of training data, *D*_*train*_.

For Period 1 we trained four 7-class models (control gestures and Rest) on sessions 1-2, 1-4, 1-6, and 1-8 (four sets of *D*_*train*_), and tested them on sessions 3-10, 5-10, 7-10, and 9-10 (four sets of *D*_*test*_), respectively (Supplementary Fig. 4a, Models 1-4, blue cells). Similarly, for Period 2, we trained four 7-class models on sessions 11-12, 11-14, 11-16, and 11-18 (four sets of *D*_*train*_), and tested them on sessions 13-20, 15-20, 17-20, and 19-20 (four sets of *D*_*test*_), respectively (Supplementary Fig. 4a, Models 5-8, pink cells). Once a model was trained, its weights were held fixed for simulating performance on the testing sessions.

To select the gestures for simulated control, we developed a class-selection algorithm that evaluated the performance of all gestures in the validation phase using the output of 14-class cross-validation models (all 13 gestures and Rest). The control gestures were selected as the six highest-performing gestures (for details, see Supplementary Method 2, Supplementary Fig. 5). Then for each set of training data, *D*_*train*_, we trained a 7-class model (six control gestures and Rest) to test on the remaining unseen data, *D*_*test*_. None of the data for testing was ever used in model training or validation. *D*_*test*_. All five blocks from a particular session were designated exclusively either for model training or testing. To simulate practical use of this training procedure, and to account for the possibility of longitudinal decline in signal quality, models were always tested on data collected following the training set.

We simulated online testing on each set of held-out testing data, *D*_*test*_, using the 7-class model (six control classes and Rest) that was trained on data from the corresponding *D*_*train*_ (see *Label assignment* for how labels were assigned to each sample). The training dataset was balanced to contain an equal number of samples per class by down-sampling all classes (including Rest) to match the class with the fewest samples.

As described above, output probabilities were computed from a 256 ms sliding window (shifted by 100 ms increments) and the smoothed probabilities were calculated (Fig. 1a-d). Once the smoothed probability of the Rest class fell below a preset threshold of 0.2 (chosen heuristically), the gesture with the highest probability was chosen (Fig. 1e) and the time stamp of the classification was stored for performance evaluation. After each classification, a refractory period of 3 s was enforced to prevent multiple classifications being produced by the same attempted gesture (Fig. 1e). The performance of each control gesture during simulated online testing was evaluated according to its sensitivity, accuracy, FPF, and latency (see Supplementary Method 3). Because *D*_*test*_ contained all 16 gestures, (including facial gestures) we excluded from our analysis any classification that occurred as a result of an attempted non-control gesture (excluded gesture).

**Figure 1.**
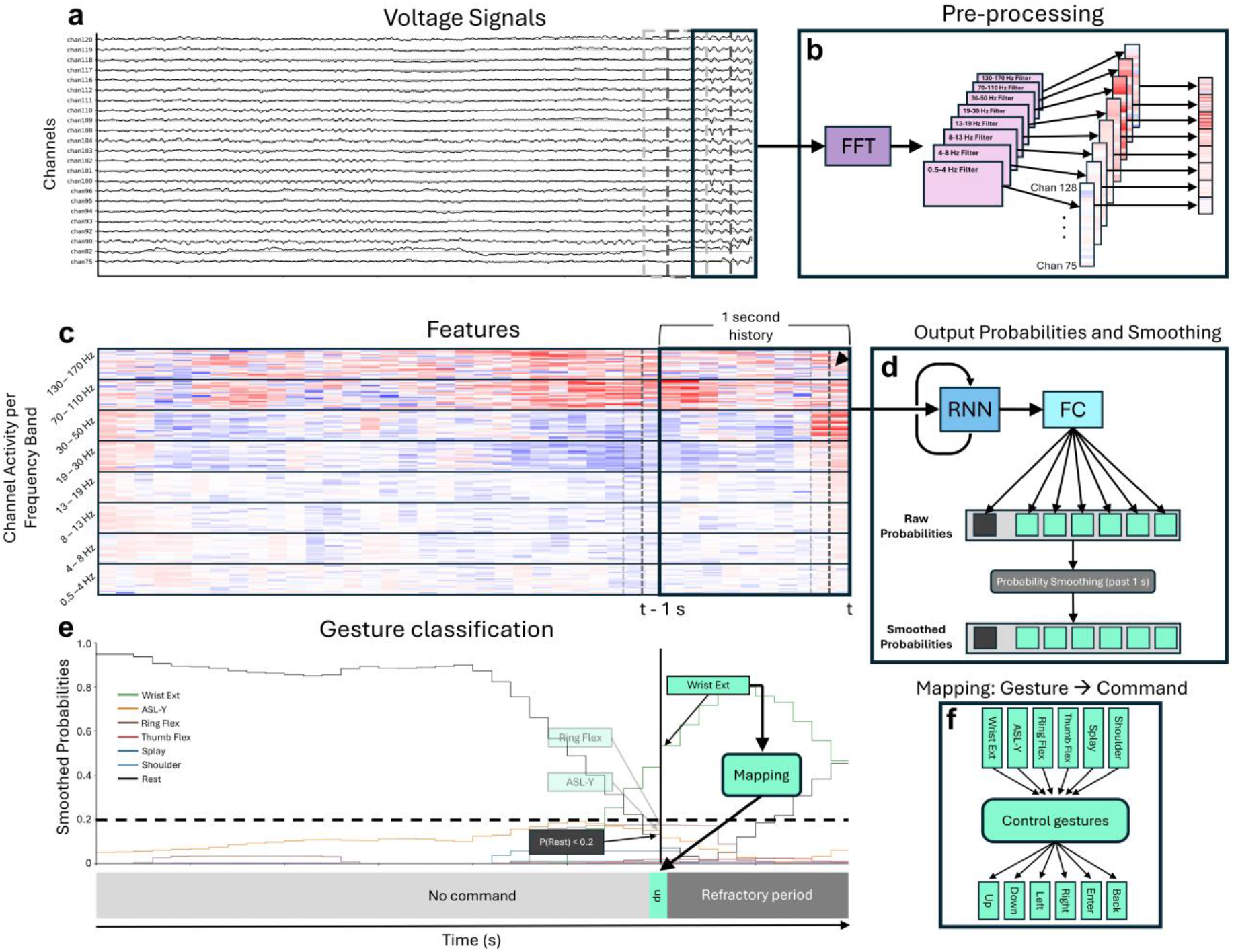
Gesture classification pipeline. (**a**) 256 ms sliding windows of voltage signals were continuously shifted by 100 ms increments. A sample from 20 representative channels is shown. (**b**) A Fast Fourier Transform (FFT) was used to compute the spectral power of these sliding windows from which the log-powers of multiple frequency bands were computed and vertically concatenated. The resulting array was placed into a 1 s running buffer (10 feature vectors computed every 100 ms). (**c**) The running buffer was then used as time history for the recurrent neural network (RNN). All possible frequency band features are displayed, though not all were used (see *Frequency band selection*). The same 20 representative channels shown in (**a**) are shown within each frequency range. (**d**) Two fully connected (FC) layers were connected to the RNN, and output class probabilities, issued every 100 ms, were smoothed using the past 1 s. The boxes in green represent the probabilities of the control gestures. The dark grey box represents the probability of Rest. (**e**) Upon the smoothed probability of the Rest class falling underneath a predetermined threshold of 0.2, the control gesture with the highest probability was selected. A possible control strategy that maps “Wrist Ext” to an “Up” command is illustrated. (**f**) A possible mapping of control gestures to UI commands.

### Label assignment

In order to train models for validation and testing, we assigned labels to each sample of the training data, *D*_*train*_. For each set of training data, *D*_*train*_, we assigned Rest and gesture labels to each time sample according to changes in the γ_high1_ power. We chose the γ_high1_ frequency band due to its rapid timescale of modulation during attempted movements^25^. We chose not to assign gesture labels based on attempted movement onset and offset. This was done to simulate the lack of ground truth for attempted movements that was expected in Locked-in Syndrome (LIS). We repeated the following label assignment process for each gesture:

1. For each channel, we concatenated segments of cue-aligned γ_high1_ power across all trials, where each trial segment was composed of all samples from −2 to 5 s relative to the onset of the visual gesture cue (Supplementary Fig. 6a, b). To account for the inter-trial variability of response latencies, we realigned the γ_high1_ power segments across all trials using a shift-warping model^26^ trained on the cue-aligned γ_high1_ power segments (Supplementary Fig. 6c, d). This model was trained on all channels that had significant peak modulation in the γ_high1_ band (see *Frequency band selection*), and realignment was performed on each trial using a trial-specific temporal shift (t_shift_[tr], Supplementary Fig. 6d-f). Note that no temporal warping (stretching or contracting) was performed in this study.
2. Using these realigned trials, we then computed the mean γ_high1_ power across all trials and all channels with significant peak modulation (Supplementary Fig. 7a).
3. We then determined the two time points relative to the onset of the visual cue where this mean γ_high1_ power crossed a threshold of 70% of the peak value (Supplementary Note 3). We designated the time point where the mean γ_high1_ power crossed above the threshold as t_gesture label onset_ and the time point where the mean γ_high1_ power returned beneath the threshold as t_gesture label offset_.
4. For each trial of a particular gesture, we assigned labels to samples between and including t_gesture label onset_ + t_shift_[tr] and t_gesture label offset_ + t_shift_[tr] relative to the onset of the visual cue (Supplementary Fig. 7b). Note that it was necessary to include the term t_shift_[tr] to the bounds where gesture labels were assigned to account for the shift that was applied to each trial.

After all gesture labels were applied, the remaining samples were labeled as Rest. Note that the primary function of the onset and offset times of the gesture labels were used to create training datasets with little overlap between Rest and gesture features, and not to define the movement onset (Supplementary Fig. 8).

### Mean β_2_ activity onset as a proxy for movement onset

It was necessary to approximate the attempted movement onset times for 1) defining the detection window which was used to determine whether gesture classifications were true or false positives and 2) computing the model latency to gesture classification (Supplementary Method 3, eqs. S8, S9). For each gesture, we simulated a lack of ground truth movement onset that was expected in LIS by using the onset of the neural activity as a proxy. Specifically, we used the β_2_ frequency band due to its tight correlation with the movement planning phase^27^, which likely biased our approximated onset times to slightly before true movement onset. For Periods 1 and 2, we used the neural activity from all trials in *D*^1^ and *D*^2^, respectively, to visually determine the onset of the mean β_2_ modulation (i.e., the event-related desynchronization or decrease in power) relative to the start of the visual cue and designated this time point as t_gesture β2 onset_ (Supplementary Fig. 9a, b). The modulation onset times for Periods 1 and 2 are reported in Supplementary Fig. 9c and did not change between the two periods by more than 0.1 s for any gesture.

### Performance metrics

We evaluated model performance during cross-validation and during testing using sensitivity, accuracy, precision, false positive frequency (FPF), and latency. (Supplementary Method 3). These metrics were computed for all gestures together and individually.

In the cross-validation phase, we computed a performance score (eq. 1) for each gesture *i* and selected our control gestures according to the gestures with the highest performance scores.

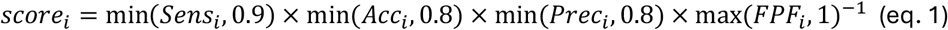

Here, we used the min() and max() functions so that the performance score for any particular gesture was not unreasonably influenced by any individual metric (Supplementary Note 4). To evaluate model performance during testing, we evaluated model sensitivity, accuracy, FPF, and latency across all control gestures together as well as for each control gesture individually.

### Model architecture and training

For each set of training data *D*_*train*_, we used a recurrent neural network (RNN) for outputting the probabilities of Rest or one of the gestures. We used this architecture to leverage the temporal patterns in neural activity.

We designed an RNN in a many-to-one architecture to capture temporal patterns in multi-band power across 1-second sequences, with labels corresponding to the sequence’s leading edge (Supplementary Fig. 10). The *b*\*60-channel multi-band power vectors were processed through an LSTM layer containing 25 hidden units for modeling sequential relationships. The output then passed through a fully-connected layer with 10 hidden units. For the cross-validation models, the output afterward passed through a final layer with 14 hidden units (13 gestures + Rest) because the performance scores of all 13 gestures needed to be evaluated (Supplementary Fig. 10a). For the models used for testing, the output instead passed through a final layer with 7 hidden units (6 control gestures + Rest) because only 6 control gestures were evaluated (Supplementary Fig. 10b). These layers used eLU and softmax activation functions respectively, with the softmax layer producing normalized probability outputs.

We trained each model through cross-validation, continuing for 15 epochs (complete forward and backward passes through the training data) until validation accuracy stabilized. We similarly trained the models for testing for 15 epochs. The loss between true and predicted labels was computed using categorical cross-entropy for batches of 45 samples. The network weights were updated using the Adam optimizer^28^. To maintain generalizability, we implemented 30% dropout regularization in both the LSTM and fully connected layers. Initial weights followed the He Normal distribution. The neural network was built using Keras running on TensorFlow 2.8.0 in Python 3.8.

### Model evaluation metrics

#### Wald test for determining model-specific performance trends

To investigate the stability of models that were used for testing, we performed a two-sided Wald test to determine whether there was a significant change (α = 0.05) over time in the slope for each performance metric. The Wald test was performed for each performance metric of models 1, 2, 3, 5, 6, and 7, which were tested on sessions 3-10, 5-10, 7-10, 13-20, 15-20, and 17-20, respectively (Supplementary Fig. 4a). The Wald test was not performed on performance metrics from models 4 and 8 because these models were only tested on two following sessions within Period 1 and Period 2 (sessions 9 and10, and 19 and 20, respectively), producing insufficient data points for the Wald test.

Additionally, we investigated whether each performance metric improved by using models trained on additional training sessions. To do this, we performed a two-sided Wald test to determine if there was a significant change (α = 0.05) in the slope of each performance metric as a function of the number of sessions that the corresponding model was trained on. Specifically, for each performance metric, our independent variable was the number of training sessions (2, 4, 6, and 8 training sessions), while our dependent variable was the mean value of the performance metric over the corresponding testing sessions (8, 6, 4, and 2 testing sessions, respectively).

#### Overall performance in Period 1 and Period 2

When comparing performance metrics of models trained and tested on data from Period 1 to metrics of models trained and tested on data from Period 2 we used a non-parametric two-sided Wilcoxon Rank-Sum test to determine whether there were statistically significant differences.

#### Composite performance metrics

For each set of testing sessions, we computed the composite performance metrics, which consisted of the composite sensitivity, composite accuracy, composite confusion matrix, and composite false positive frequency (FPF). For each set of testing sessions, these composite performance metrics represented the overall sensitivity, accuracy, confusion matrix, and FPF across all the sessions in that testing set. For each testing set, the composite sensitivity and accuracy were computed by taking the mean sensitivity and accuracy, respectively, across all sessions in that testing set. We then computed the composite sensitivity and accuracy for each gesture. Additionally, for each testing set, we computed the composite confusion matrix, which represented the element-wise sum of all individual confusion matrices in that testing set. Finally, for each testing set the composite FPF was computed by taking the mean of all FPFs across all testing sessions in that testing set.

### Signal evaluation

#### Comparing modulation amplitude

We compared the modulation amplitudes of each channel between Periods 1 and 2 across all control gestures that were selected at least once in each period and across all frequency bands with significant modulation (for details, see Supplementary Fig. 11). For each channel, we compared trial means of the Periods 1 and 2 using a Welch’s t-test. To account for multiple comparisons (1 t-test per channel = 60 comparisons), we performed a false discovery rate correction (q = 0.05).

#### Between-class separation

We then compared the between-class separation in neural features between Period 1 and Period 2. We investigated the neural features from each frequency band with significant modulation individually (60 dimensions) or altogether (*b*\*60 dimensions). First, for each gesture, we realigned all 200 trials from Period 1 and all 200 trials from Period 2 using only γ_high1_-based shift-warping models and partitioned the realigned trials from each period into contiguous sets of 40 trials/gesture (trials from two contiguous sessions). This was so that we could create a sample of multiple distance metrics for significance testing. For each contiguous set, we then computed each trial’s mean activity and projected it into a low dimensional manifold by 1) reducing the dimensionality of the data using principal component analysis (PCA), and 2) optimally separating the classes using linear discriminant analysis (LDA).

1. For each contiguous set, we reduced the dimensionality from either 60 or *b*\*60 dimensions to the number of principal components that explained 90% of the variance in the data. Due to the relatively small number of trials compared to the number of features, we applied PCA before LDA to prevent the linear discriminant model from overfitting to irrelevant features (i.e., features with no or low predictive power and random noise).
2. We then reduced these components to a (C-1)-dimensional manifold using LDA, where C was the number of gestures that were compared. C-1 is the largest possible dimensionality of the manifold space when using LDA. This manifold space accounted for 99 - 100% of the explained variance in the PC-transformed data computed in 1). We used LDA to improve between-class separation because it maximizes the ratio of between-class scatter to within-class scatter (i.e., it maximizes the separation between class means while minimizing the spread within each class).

Finally, we computed the Mahalanobis distance (*d*, eq. 2) between the manifold representations for each pair of control gestures.

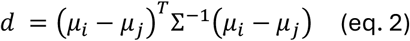

where *μ*_*i*_ and *μ*_*j*_ are the means of gestures *i* and *j*, respectively, in the manifold space and Σ is the pooled covariance matrix of the distributions of gestures *i* and *j*. In this case, Σ is the mean of the covariance matrices of *i* and *j* because each distribution has an equal number of samples (eq. 3):

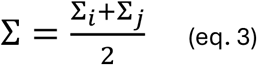

## Results

### Selection of frequency bands and control gestures

We excluded the δ (delta, 0.5 - 4 Hz), θ (theta, 4 - 8 Hz), and μ (Mu, 8 - 13 Hz) frequency bands from this analysis as they did not contain any significant gesture-specific modulation (for each gesture, the nadir of the trial-averaged z-score was not less than −1.96). However, modulation was visible in the spectral analysis for all three bands to some degree. The remainder of the results were evaluated using the β_1_ (beta 1, 13 - 19 Hz), β_2_ (beta 2, 19 - 30 Hz), γ_low_ (low gamma, 30 - 50 Hz), γ_high1_ (high gamma 1, 70 - 110 Hz), and γ_high2_ (high gamma 2, 130 - 170 Hz) frequency bands as these did contain significant gesture-specific modulation (for at least one gesture, the nadir of the trial-averaged z-score was less than −1.96 for β_1_, β_2_, γ_low_ and the peak of the trial-averaged z-score greater than 1.96 for γ_high1_ and γ_high2_).

The control gestures that were selected for models trained on data from Period 1 or on data from Period 2 are shown in Supplementary Fig. 4b. For each model, these were the gestures with the highest performance scores in the validation phase. For models trained on data from Period 1 the control gestures that were most frequently selected were Wrist Ext (selected for 4/4 models), Ring Flex (4/4 models), Thumb Flex (4/4 models), ASL-Y (4/4 models), Splay (2/4 models), Wrist Flex (2/4 models), Shoulder (2/4 models), Pinch (1/4 models), and Grasp (1/4 models). For models trained on data from Period 2 the control gestures that were most frequently selected were Wrist Ext (selected for 4/4 models), Splay (4/4 models), ASL-Y (3/4 models), Shoulder (3/4 models), Wrist Flex (3/4 models), Middle Flex (3/4 models), Thumb Flex (2/4 models), Ring Flex (1/4 models), Grasp (1/4 models), Pinky Flex (1/4 models).

### Model performance across Period 1 and Period 2

We evaluated the simulated performance of models trained and tested on data from Period 1 and models trained and tested on data from Period 2 (Fig. 2, Supplementary Tables 2, 3). Within Period 1 and Period 2 performance metrics of all the models generally remained stable except for the accuracy of Model 3, trained on sessions 1-6 (Fig. 2d), which showed a significant decline of −0.54% per day (*P* = 0.03; two-sided Wald test). We also evaluated the simulated performance of the models as a function of the number of sessions used for model training. The accuracy of Models 5-8 (tested on data from Period 2) showed a significant increase of 2.9% per additional training session (*P* = 0.03; two-sided Wald test) where Models 5-8 trained on 40 - 160 trials per control gesture, respectively. No other performance metrics showed significant changes with respect to the number of training sessions for either Period 1 or Period 2.

**Figure 2.**
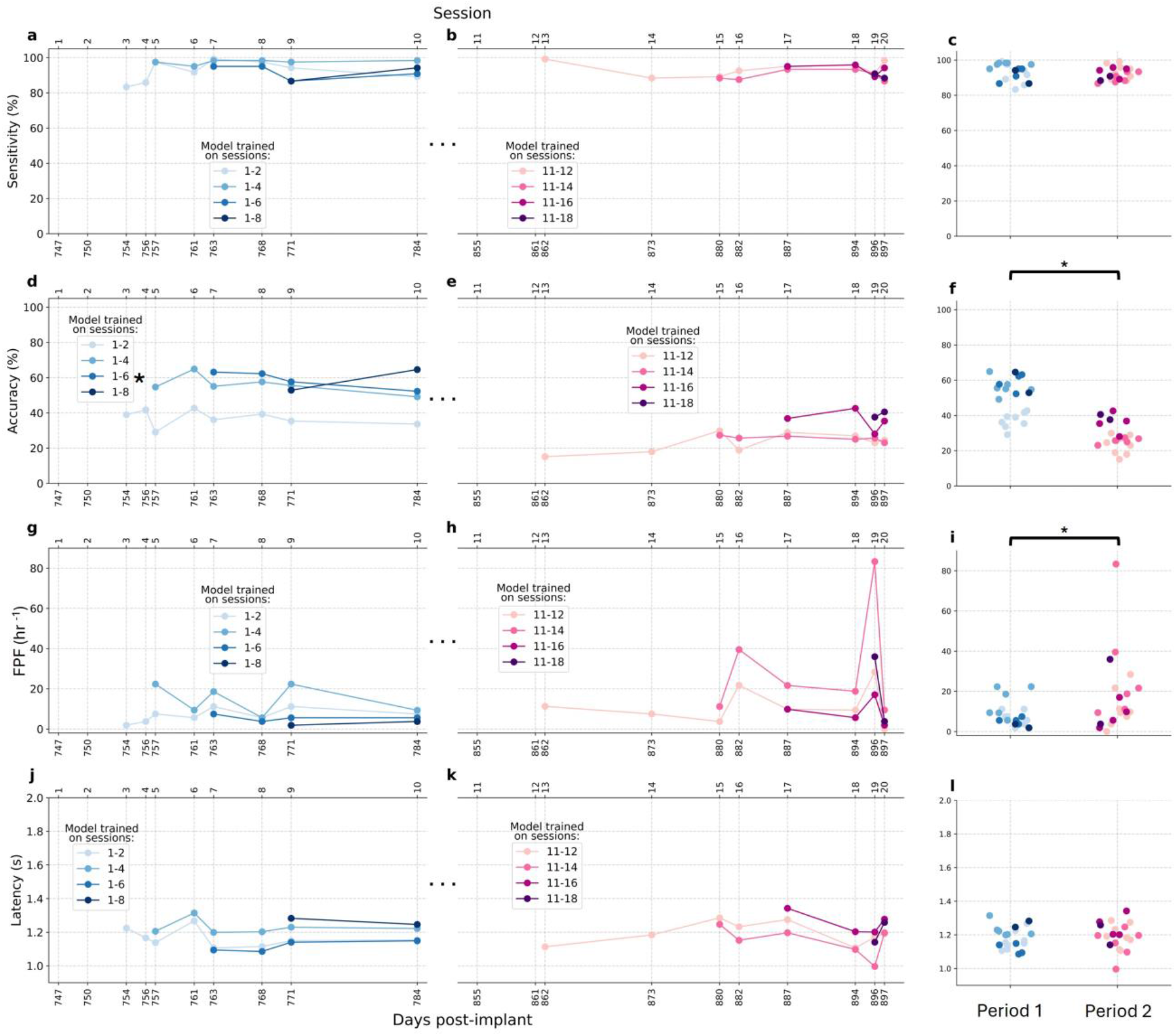
Offline performance. Sensitivity, accuracy, FPF, and latency of models trained and tested using data exclusively from Period 1 (**a, d, g**, and **j**, respectively) and exclusively from Period 2 (**b, e, h**, and **k**, respectively). Performance metrics of models trained on more data (and consequently tested on less data) are shown in darker colors. Data collected during days 747 and 750 were used only for model training and consequently do not have performance metrics. Performance metrics with a significant change over time are represented by a * (*P* < 0.05). For each subplot, the bottom horizontal axis represents the number of days post-implant on which the data was collected, and the top horizontal axis represents the corresponding session numbers. The ellipses between days 784 and 855 represent a gap of 71 days in data collection for this task. Note that the session numbers do not represent the overall number of recording sessions with the participant during the clinical trial. **c, f, i**, and **l** represent the corresponding summary distributions of the sensitivities, accuracies, FPFs, and latencies, respectively, and significant differences between performance metrics are represented by a * (*P* < 0.05).

Between Period 1 and Period 2, performance metrics either remained stable or became worse. The mean model sensitivity and latency remained stable between Period 1 and Period 2 (93.6% compared to 92.1% and 1.18 s compared to 1.19 s, respectively) (Fig. 2c, l). However, the classification accuracy decreased from 49.3% for Period 1 to 28.0% in Period 2 (*P* = 2.1 × 10^−6^; Fig. 3f), and the false positive frequency (FPF) increased from 8.5 hr^−1^ for Period 1 to 17.5 hr^−1^ for Period 2 (*P* = 3.9 × 10^−2^; Fig. 2i). Additionally, for each set of testing sessions, the composite performance metrics are shown in Supplementary Fig. 12 (sensitivity, accuracy, and confusion matrix) and in Supplementary Fig. 13 (FPF).

**Figure 3.**
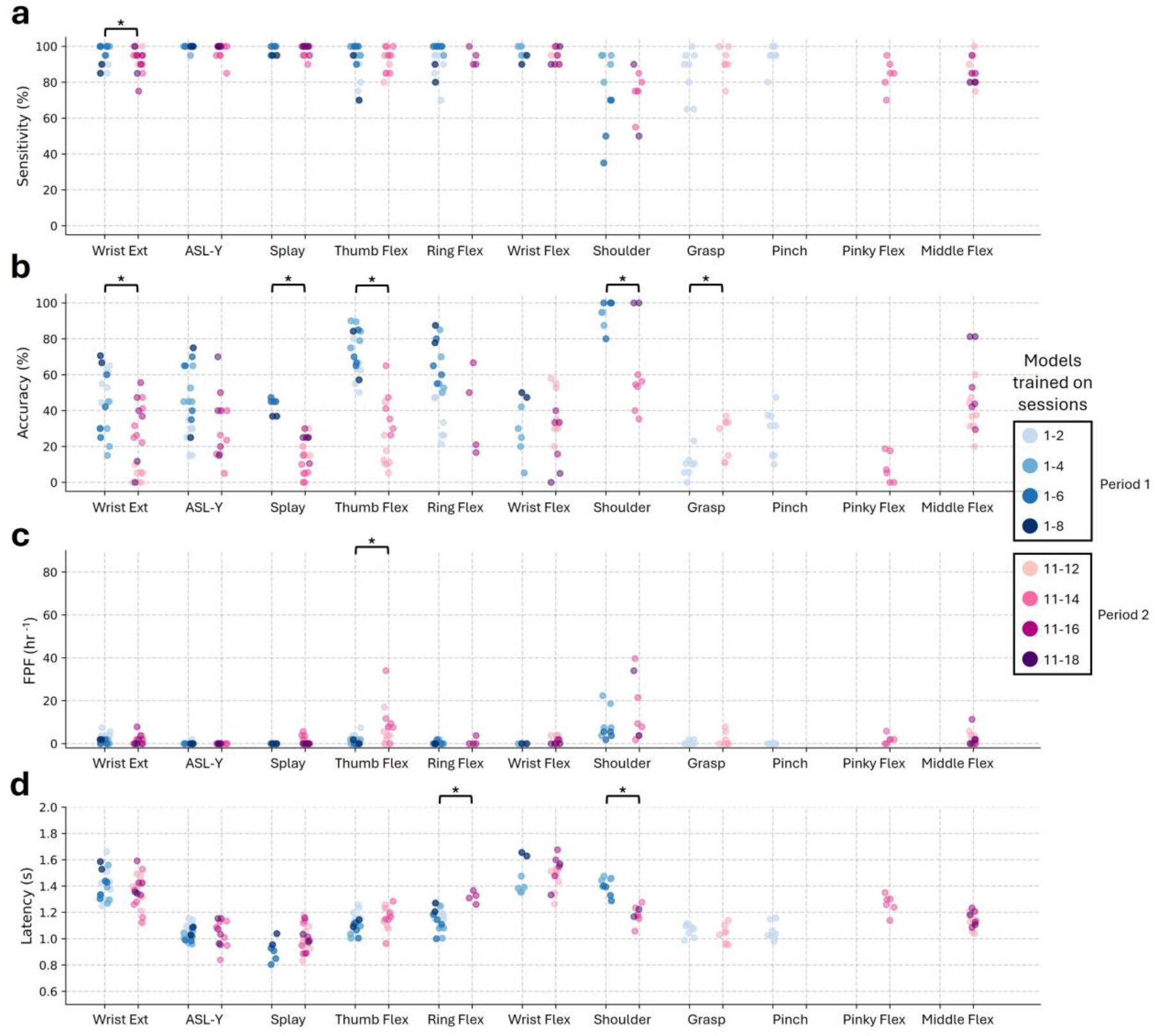
Offline performance metrics per gesture. Sensitivity (**a**), accuracy (**b**), false positive frequency (**c**), and latency (**d**) of each gesture for models trained and tested using data exclusively from Period 1 and for models trained and tested using data exclusively from Period 2. Significant differences between performance metrics are represented by a * (*P* < 0.05).

We then assessed the performance of each of the control gestures that were determined for models trained and tested on data from Period 1 and for models trained and tested on data from Period 2 (Fig. 3). We compared performance metrics of the control gestures that were selected at least once across both types of models. Specifically, these were Wrist Ext (selected for 8/8 models in total), ASL-Y (7/11 models), Splay (6/8 models), Thumb Flex (6/8 models), Ring Flex (5/8 models), Wrist Flex (5/8 models), Shoulder (4/8 models), and Grasp (2/8 models). Sensitivity, FPF, and latency generally remained similar across Periods 1 and 2 for most gestures (Fig. 3a, d, Supplementary Table 4a, d). Note that most of the false positives in Period 2 were caused by Thumb Flex (7.9 hr^−1^) and Shoulder (15.2 hr^−1^), and the false positive frequency (FPF) for Thumb Flex was significantly higher in Period 2 compared to Period 1 (Fig. 3c, Supplementary Table 4c). However, there were more gestures with changes in classification accuracy than changes in any other metric. The classification accuracies for Wrist Ext, Splay, Thumb Flex, and Shoulder all decreased from Period 1 to Period 2 by 25.6 - 44.4% (Fig. 3b, Supplementary Table 4b).

### Changes in modulation and between-class separation across Period 1 and Period 2

To investigate possible causes for changes in our performance metrics, we first assessed whether there was a change in the modulation of any of the frequency bands (β_1_, β_2_, γ_low_, γ_high1_, and γ_high2_) from Period 1 to Period 2 (Fig. 4). We compared each channel’s change in modulation for each of the control gestures that were selected at least once across both periods (Fig. 4a).

**Figure 4.**
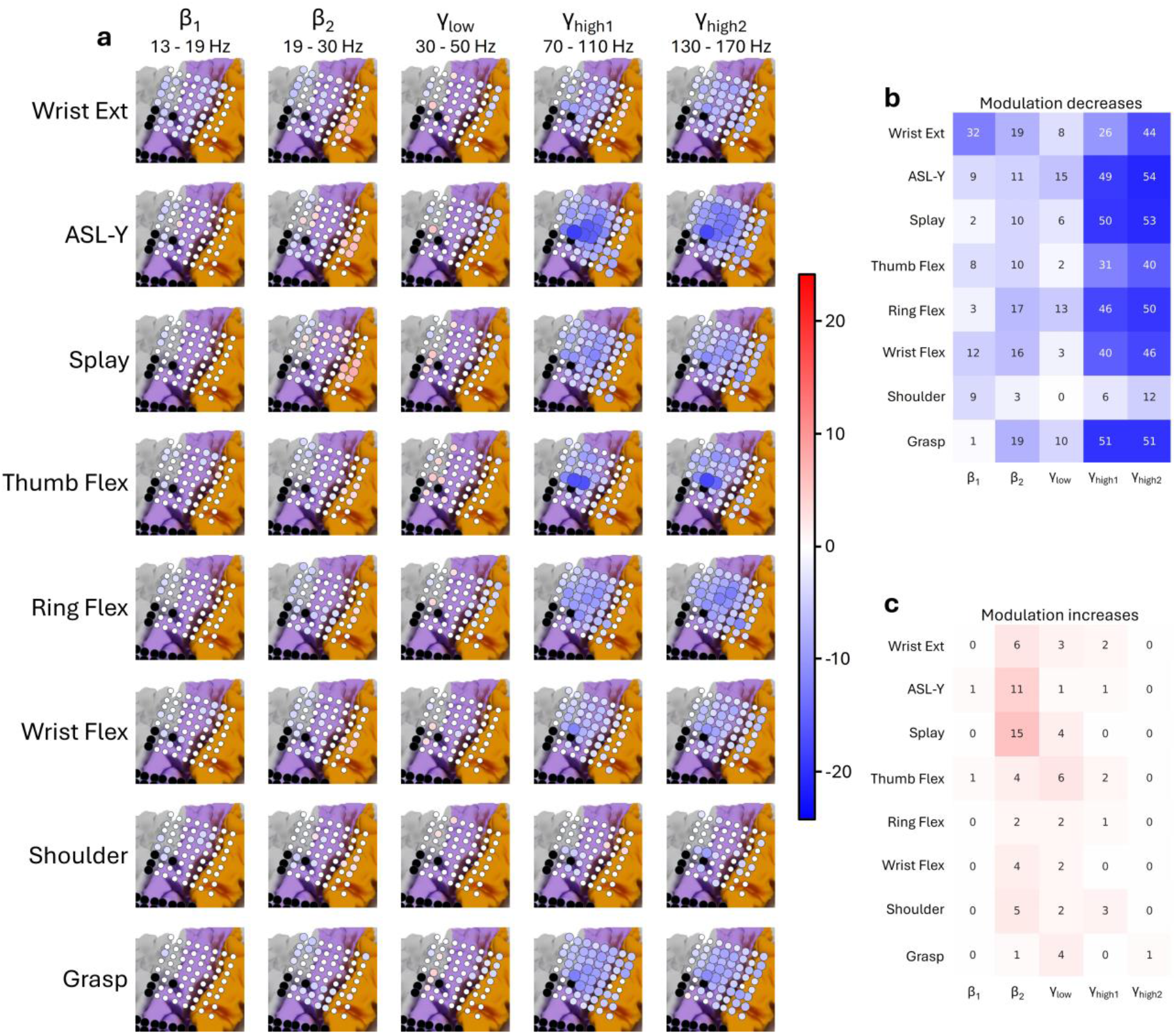
Changes in modulation between Period 1 and Period 2. (**a**) For each frequency band and gesture, the changes in modulation between Period 1 and Period 2 across all channels in the dorsal grid are shown and overlayed on a schematic of the left hemisphere. Channels overlayed with red or blue markers indicate a significant increase or decrease in modulation, respectively (*P* < 0.05; t-test, Bonferroni corrected). For each channel, the change in modulation is represented by the corresponding positive or negative t-statistic. Markers are darker and larger if the t-statistics are greater in magnitude. Channels for which there was no significant change in modulation are marked in white. Channels marked in black were not used in this study. Color coding of the cortex indicates anatomical landmarks: magenta for pre-central gyrus and orange for post-central gyrus. For each frequency band and gesture, (**b**) and (**c**) represent the total number of channels with significant decreases and increases in modulation between Period 1 and Period 2, respectively.

A drop in modulation was observed across all frequency bands for all attempted upper limb gestures but was especially prominent in the γ_high1_ and γ_high2_ frequency bands (Fig 4a, Supplementary Figs. 14-21). For all movements, a drop in γ_high1_ or γ_high2_ band power modulation was observed in 6 - 54 channels (median of 46 channels, Fig. 4b). Drops in modulation for ASL-Y and Thumb Flex were especially pronounced and were highly concentrated in the cortical hand knob region of motor cortex. For example, the γ_high1_ and γ_high2_ modulation of channel 91 dropped by 52% and 68%, respectively, for ASL-Y (Supplementary Fig. 22). Increases in modulation were sparse and were primarily observed in the lower frequency bands (Fig. 4c).

We then assessed the separation between classes by computing the average Mahalanobis distance (*d*) between pairs of control gestures that were selected at least once across both periods.

Using all frequency bands, the average Mahalanobis distance, *d*, for Period 1 ranged between 3.90 - 9.16. For Period 2, the average *d* of almost all gesture pairings significantly dropped to a range of 3.23 - 6.69 (Fig. 5a). To visualize these differences, we plotted these features in a neural manifold space (Fig. 5b). Moreover, the largest Mahalanobis distances for Period 1 were between attempted Shoulder movement and all other attempted gestures (ASL-Y, Wrist Flex, Splay, Wrist Ext, Thumb Flex, Ring Flex, Grasp). This separation of attempted Shoulder movements remained relatively large in Period 2 and could have been due to modulation that was more concentrated in the upper right portion of the ECoG grid (Supplementary Fig. 20) compared to other attempted gestures. Further, this may also explain the higher accuracy of attempted Shoulder movements across testing sessions compared to other gestures (Fig. 3b, Supplementary Table 4b).

**Figure 5.**
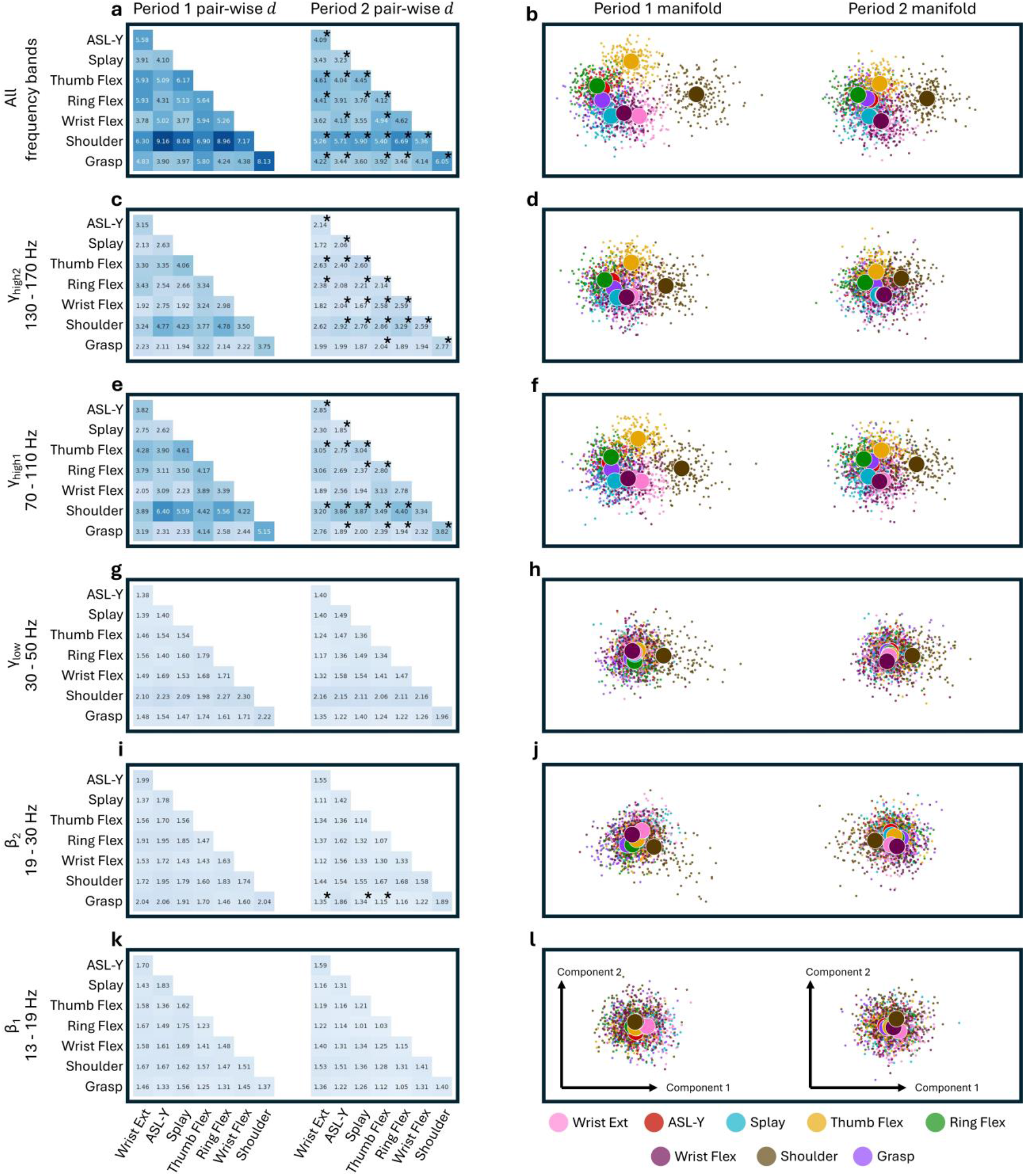
Changes in gesture separation between Period 1 and Period 2. For all frequency bands combined, and for each individual frequency band, the changes in gesture separation are quantified as Mahalanobis distances (*d*) and represented in a low-dimensional space. (**a, c, e, g, i, k**) Matrices of average *d* between pairs of control gestures using data from Period 1 (left triangular matrices) and data from Period 2 (right triangular matrices). For any pair of control gestures, significant differences in the average *d* between Period 1 and Period 2 are represented by a * (*P* < 0.05) in the Period 2 matrices. (**b, d, f, h, j, l**) Low dimensional visualization of control gestures in the manifold space. The centroids of each attempted gesture are marked by large circles. The space was created by using the first two principal components of the linear discriminant reduced space.

We then investigated the changes in the Mahalanobis distance, *d*, for each frequency band. The drops in the average *d* from Period 1 to Period 2 occurred mostly in the γ_high1_ and γ_high2_ frequency bands. For γ_high2_, the average *d* of almost all gesture pairings significantly dropped from 2.14 - 4.78 to 1.89 - 3.29 (Fig. 5c). For γ_high1_, the average *d* of many gesture pairings significantly dropped from 2.31 - 6.40 to 1.67 - 4.40 (Fig. 5e). The drops in the average *d* were clearly visible as the between-class clustering was tighter in the neural manifold space (Fig. 5d, f). The distances of gesture pairings in the β_1_, β_2_, and γ_low_ frequency bands were lower relative to the γ_high1_ and γ_high2_ frequency bands (1.23 - 2.30, 1.37 - 2.06, and 1.23 - 1.75, respectively) and did not substantially change from Period 1 to Period 2, aside from three gesture pairings in the β_2_ frequency band (Fig. 5g-l).

## Discussion

Gesture-based decoding has been explored with able-bodied clinical epilepsy patients^29–32^ and more recently, with BCI clinical trial participants who had severe movement impairments from spinal cord injury^22,33–37^ or brainstem stroke^24,38^. However, unlike these conditions, amyotrophic lateral sclerosis (ALS) has an inherent neurodegenerative progression^1^. Despite previous work with gesture-based decoders for people with ALS^11,22,36^, a months-long evaluation of such a decoder with this changing disease state has not yet been reported. Here we investigated the decoding potential of attempted upper limb gestures from ECoG signals in a clinical trial participant with ALS within and across two time periods separated by 5 months.

We recorded a variety of attempted upper limb, lower limb, and facial gestures because we did not have an a priori set of classes designed for multiclass decoding. We therefore developed a class-selection algorithm that systematically selected a set of optimal control gestures by estimating their performance during validation with the same metrics that were assessed during testing.

Because such a class-selection algorithm was based on the same metrics used for testing, we believe it was superior to selecting classes based on visualizing or quantifying class separation, which becomes increasingly difficult with larger numbers of classes. Additionally, such a class-selection algorithm can significantly save time in developing multiclass classifiers, as different subsets of classes would not have to be laboriously recorded, tested, and compared.

For both Period 1 and Period 2, we trained models on data from various numbers of sessions and tested them on data from held out “unseen” sessions. Performance metrics of the trained models generally did not change over time (aside from the classification accuracy of Model 3), suggesting that neural signals were relatively stable within Period 1 and within Period 2. However, it may be that none of the testing sets spanned a long enough period of time for performance declines (such as the decline in Model 3’s accuracy) to be observed in other models. Though the accuracy of models trained and tested on data from Period 2 improved with increasing training sessions (Fig. 2e), accuracy comparable to Period 1 was not recovered. Conversely, the lack of significant improvement in accuracy with increasing training sessions during Period 1 could suggest that neural representations in that period may have been more stable than in Period 2, requiring less training sessions to achieve high testing accuracy (Fig. 2d).

Overall, the accuracy of control gesture classification was higher in Period 1 compared to the later Period 2 (Fig. 2d-f). The lower classification accuracy in Period 2 was likely related to the observed reduction in high gamma band power modulation (Fig. 4) and reduced between-class separation (Fig. 5). The reduction in γ_high1_ and γ_high2_ modulation was especially pronounced for attempted ASL-Y and Thumb Flex gestures with the greatest drops in modulation occurring in channels 91 and 92, located over the cortical “Hand knob” region. Our findings are consistent with previous work reporting longitudinal power decreases in similar frequency bands from a participant with ALS^20^. Moreover, the lower classification accuracy was likely related to a reduction in the Mahalanobis distances across pairs of gesture representations, especially within the γ_high1_ and γ_high2_ frequency bands (Figs. 5c-f). The neural representations could have shifted toward a weaker but more uniform activity profile, thereby increasing gesture similarity and decreasing between-gesture separability. The between-gesture separation among the lower frequency bands remained poor, consistent with previous work demonstrating poor classification accuracy using only low frequency power^39^. On the other hand, sensitivity remained high in both Period 1 and Period 2, but false positive frequency (FPF) increased in Period 2. However, this may in part have been due to our gesture detection threshold of 0.2, which was high enough to allow for true-positive detections, but also too high to filter out false positive detections.

To our knowledge this is the first study reporting on gesture decoding over a months-long timescale for a clinical trial participant with ALS. However, as this is an N-of-1 study, our results may be specific to our clinical trial participant and must be verified by further work. A limitation of this study is that since decoding was simulated (albeit with the same processing pipeline that would have been used online), the participant had not benefited from closed-loop visual feedback.

Previous studies have indicated that neural representations of facial and upper limb and facial gestures become more separable with repeated closed loop trials^38,40^.

The longitudinal decline we observed in BCI performance, high gamma modulation, and between-gesture separability were not accompanied by a significant deterioration in our participant’s neurological exam. The MRC scale that was used to code for muscle strength in periodic neurological examinations is a somewhat coarse categorical measure that may not capture subtle changes in strength over time. Our participant could not grip a dynamometer to obtain a more continuous measure of strength. Nevertheless, our participant denied any significant subjective decline in strength during the 5-month period covered by this study, and we did not observe any decline in his effort during experimental sessions. Notwithstanding these methodological limitations, our observations raise the possibility that decoding performance and signal modulation and separability in an ECoG BCI can decline in users with ALS without any observable decline in strength.

ECoG BCI performance in this and other studies^13–15,20,24,41^ has relied in large part on modulation of local field potential activity in high gamma frequencies. Electrophysiological studies in animal models have indicated that broadband high gamma activity likely reflects changes in the aggregate firing rates of local neural populations in proximity to recording electrodes^25^. With progressive neuronal loss due to ALS, this population activity may also be expected to decline. Thus, modulation of high gamma activity and resulting BCI performance could represent a potential biomarker for disease progression in the absence of overt changes in neurological signs and symptoms. Further research is needed to test this hypothesis, but if true, this could potentially provide a more immediate and sensitive measure of the efficacy of new therapeutics for ALS and other progressive degenerative neurological disorders.

## Data availability

Source data for rendering the figures in the main text and supplementary information is publicly available at [*link will be available after manuscript acceptance*]. Beginning immediately after publication, neural data from the study participant and the study protocol will be available from the corresponding author upon reasonable request.

## Code availability

The analytical code for rendering the figures in the main text and supplementary information is publicly available at [*link will be available after manuscript acceptance*]. Code used for offline model development and post-hoc analysis is also included. Model training and offline analysis were done using Python (version 3.9.13). The recurrent neural network was built using Keras with a TensorFlow backend (version 2.8.0).

## Acknowledgements

The authors wish to express their sincere gratitude to participant CC01, whose dedication and efforts made this study possible. The authors thank William Anderson and Chad Gordon for medical and surgical support during device implantation; the ALS clinic at the Johns Hopkins Hospital and the Johns Hopkins ALS Clinical Trials Unit for their consultations on ALS-related topics and care of the participant; Kevin Nathan and Mathijs Raemaekers for analysis of fMRI data; Yujing Wang for 3D reconstruction of the brain MRI and electrode co-registration with the post-operative CT; Brock Wester for help with illustrations.

## Contributions

D.N.C., M.S.F., and N.E.C. conceived and designed the experiments. D.N.C. designed and implemented the detection algorithm and the neural classifier. D.N.C., M.A., and S.L. contributed to data analysis and interpretation. R.G. and C.C. contributed to task design. G.W.M. contributed to design of the simulated offline pipeline. M.J.V., F.V.T., N.F.R., M.S.F., and N.E.C. contributed to patient recruitment and regulatory approval. N.J.M., L.C. and A.U. contributed to the clinical care of the participant. N.E.C. and K.R.R. and planned and performed device implantation. D.N.C., S.L., F.V.T., N.F.R., and M.S.F. were involved in surgical planning and intraoperative functional mapping. N.F.R. and N.E.C. obtained funding. N.E.C. supervised the study. D.N.C., M.S.F., and N.E.C. prepared the manuscript. All authors reviewed and revised the manuscript.

## References

1. Masrori, P. & Van Damme, P. Amyotrophic lateral sclerosis: a clinical review. Euro J of Neurology 27, 1918–1929 (2020).

2. Donaghy, C., Thurtell, M. J., Pioro, E. P., Gibson, J. M. & Leigh, R. J. Eye movements in amyotrophic lateral sclerosis and its mimics: a review with illustrative cases. Journal of Neurology, Neurosurgery & Psychiatry 82, 110–116 (2011).

3. Sharma, R. Oculomotor Dysfunction in Amyotrophic Lateral Sclerosis: A Comprehensive Review. Arch Neurol 68, 857 (2011).

4. Spataro, R., Ciriacono, M., Manno, C. & La Bella, V. The eye-tracking computer device for communication in amyotrophic lateral sclerosis. Acta Neurol Scand 130, 40–45 (2014).

5. Holz, E. M., Botrel, L., Kaufmann, T. & Kübler, A. Long-Term Independent Brain-Computer Interface Home Use Improves Quality of Life of a Patient in the Locked-In State: A Case Study. Archives of Physical Medicine and Rehabilitation 96, S16–S26 (2015).

6. Käthner, I., Kübler, A. & Halder, S. Comparison of eye tracking, electrooculography and an auditory brain-computer interface for binary communication: a case study with a participant in the locked-in state. J NeuroEngineering Rehabil 12, 76 (2015).

7. Kang, B.-H., Kim, J.-I., Lim, Y.-M. & Kim, K.-K. Abnormal Oculomotor Functions in Amyotrophic Lateral Sclerosis. J Clin Neurol 14, 464 (2018).

8. Farr, E., Altonji, K. & Harvey, R. L. LOCKED‐IN Syndrome: Practical Rehabilitation Management. PM&R 13, 1418–1428 (2021).

9. Moss, H. E. et al. Cross-sectional evaluation of clinical neuro-ophthalmic abnormalities in an amyotrophic lateral sclerosis population. Journal of the Neurological Sciences 314, 97–101 (2012).

10. Vansteensel, M. J. et al. Fully Implanted Brain–Computer Interface in a Locked-In Patient with ALS. N Engl J Med 375, 2060–2066 (2016).

11. Mitchell, P. et al. Assessment of Safety of a Fully Implanted Endovascular Brain-Computer Interface for Severe Paralysis in 4 Patients: The Stentrode With Thought-Controlled Digital Switch (SWITCH) Study. JAMA Neurol 80, 270 (2023).

12. Willett, F. R. et al. A high-performance speech neuroprosthesis. Nature 620, 1031–1036 (2023).

13. Luo, S. et al. Stable Decoding from a Speech BCI Enables Control for an Individual with ALS without Recalibration for 3 Months. Advanced Science 10, 2304853 (2023).

14. Angrick, M. et al. Online speech synthesis using a chronically implanted brain–computer interface in an individual with ALS. Sci Rep 14, 9617 (2024).

15. Candrea, D. N. et al. A click-based electrocorticographic brain-computer interface enables long-term high-performance switch scan spelling. Commun Med 4, 207 (2024).

16. Pels, E. G. M. et al. Stability of a chronic implanted brain-computer interface in late-stage amyotrophic lateral sclerosis. Clinical Neurophysiology 130, 1798–1803 (2019).

17. Chao. Long-term asynchronous decoding of arm motion using electrocorticographic signals in monkey. Front.Neuroeng. (2010) doi:10.3389/fneng.2010.00003.

18. Sun, F. T., Arcot Desai, S., Tcheng, T. K. & Morrell, M. J. Changes in the electrocorticogram after implantation of intracranial electrodes in humans: The implant effect. Clinical Neurophysiology 129, 676–686 (2018).

19. Wyse-Sookoo, K. et al. Stability of ECoG high gamma signals during speech and implications for a speech BCI system in an individual with ALS: a year-long longitudinal study. J. Neural Eng. 21, 046016 (2024).

20. Vansteensel, M. J. et al. Longevity of a Brain–Computer Interface for Amyotrophic Lateral Sclerosis. N Engl J Med 391, 619–626 (2024).

21. Branco, M. P., Pels, E. G. M., Nijboer, F., Ramsey, N. F. & Vansteensel, M. J. Brain-Computer interfaces for communication: preferences of individuals with locked-in syndrome, caregivers and researchers. Disability and Rehabilitation: Assistive Technology 18, 963–973 (2023).

22. Willett, F. R. et al. Hand Knob Area of Premotor Cortex Represents the Whole Body in a Compositional Way. Cell 181, 396–409.e26 (2020).

23. Schalk, G., McFarland, D. J., Hinterberger, T., Birbaumer, N. & Wolpaw, J. R. BCI2000: A General-Purpose Brain-Computer Interface (BCI) System. IEEE Trans. Biomed. Eng. 51, 1034–1043 (2004).

24. Silversmith, D. B. et al. Plug-and-play control of a brain–computer interface through neural map stabilization. Nat Biotechnol 39, 326–335 (2021).

25. Ray, S., Crone, N. E., Niebur, E., Franaszczuk, P. J. & Hsiao, S. S. Neural Correlates of High-Gamma Oscillations (60–200 Hz) in Macaque Local Field Potentials and Their Potential Implications in Electrocorticography. J. Neurosci. 28, 11526–11536 (2008).

26. Williams, A. H. et al. Discovering Precise Temporal Patterns in Large-Scale Neural Recordings through Robust and Interpretable Time Warping. Neuron 105, 246–259.e8 (2020).

27. Engel, A. K. & Fries, P. Beta-band oscillations — signalling the status quo? Current Opinion in Neurobiology 20, 156–165 (2010).

28. Kingma, D. P. & Ba, J. Adam: A Method for Stochastic Optimization. (2014) doi:10.48550/ARXIV.1412.6980.

29. Leuthardt, E. C., Schalk, G., Wolpaw, J. R., Ojemann, J. G. & Moran, D. W. A brain–computer interface using electrocorticographic signals in humans. J. Neural Eng. 1, 63–71 (2004).

30. Schalk, G. et al. Two-dimensional movement control using electrocorticographic signals in humans. J. Neural Eng. 5, 75–84 (2008).

31. Chestek, C. A. et al. Hand posture classification using electrocorticography signals in the gamma band over human sensorimotor brain areas. Journal of Neural Engineering 10, 026002 (2013).

32. Hotson, G. et al. Individual finger control of a modular prosthetic limb using high-density electrocorticography in a human subject. Journal of Neural Engineering 13, 026017 (2016).

33. Wang, W. et al. An Electrocorticographic Brain Interface in an Individual with Tetraplegia. PLoS ONE 8, e55344 (2013).

34. Bouton, C. E. et al. Restoring cortical control of functional movement in a human with quadriplegia. Nature 533, 247–250 (2016).

35. Ajiboye, A. B. et al. Restoration of reaching and grasping movements through brain-controlled muscle stimulation in a person with tetraplegia: a proof-of-concept demonstration. The Lancet 389, 1821–1830 (2017).

36. Degenhart, A. D. et al. Remapping cortical modulation for electrocorticographic brain– computer interfaces: a somatotopy-based approach in individuals with upper-limb paralysis. J. Neural Eng. 15, 026021 (2018).

37. Handelman, D. A. et al. Shared Control of Bimanual Robotic Limbs With a Brain-Machine Interface for Self-Feeding. Front. Neurorobot. 16, 918001 (2022).

38. Natraj, N. et al. Sampling representational plasticity of simple imagined movements across days enables long-term neuroprosthetic control. Cell 188, 1208–1225.e32 (2025).

39. Jiang, T. et al. Characterization and Decoding the Spatial Patterns of Hand Extension/Flexion using High-Density ECoG. IEEE Trans. Neural Syst. Rehabil. Eng. 25, 370–379 (2017).

40. Śliwowski, M., Martin, M., Souloumiac, A., Blanchart, P. & Aksenova, T. Impact of dataset size and long-term ECoG-based BCI usage on deep learning decoders performance. Front. Hum. Neurosci. 17, 1111645 (2023).

41. Metzger, S. L. et al. A high-performance neuroprosthesis for speech decoding and avatar control. Nature 620, 1037–1046 (2023).

