## Supplemental Materials for "Longitudinal study of gesture decoding in a clinical trial participant with ALS"

### Supplementary Materials

Daniel N. Candrea[1✉], Miguel Angrick[2], Shiyu Luo[1], Rohit Ganji[2], Christopher Coogan[2], Griffin W. Milsap[3], Kathryn R. Rosenblatt[4], Alpa Uchil[2], Lora Clawson[2], Nicholas J. Maragakis[2], Mariska J. Vansteensel[5], Francesco V. Tenore[3], Nicholas F. Ramsey[5], Matthew S. Fifer[3], Nathan E. Crone[2]

✉

1 Department of Biomedical Engineering, Johns Hopkins University School of Medicine, Baltimore, MD

2 Department of Neurology, Johns Hopkins University School of Medicine, Baltimore, MD

3 Research and Exploratory Development Department, Johns Hopkins University Applied Physics Laboratory, Laurel, MD

4 Department of Anesthesiology and Critical Care Medicine, Johns Hopkins University School of Medicine, Baltimore, MD

5 Department of Neurology and Neurosurgery, UMC Utrecht Brain Center, Utrecht, The Netherlands

### Table of Contents

#### Supplementary Notes

Supplementary Note 1 | ALS functional rating scale (ALSFRS-R). Page 4

Supplementary Note 2 | Neural representations of gestures may drift. Page 7

Supplementary Note 3 | Label assignment using the 70% threshold crossing. Page 7

Supplementary Note 4 | Min() and max() function for performance score. Page 7

#### Supplementary Methods

Supplementary Method 1 | Clinical trial inclusion and exclusion criteria. Page 8

Supplementary Method 2 | Selecting control gestures for model testing. Page 10

Supplementary Method 3 | Performance Metrics. Page 11

Supplementary Method 4 | Lack of specificity and false positive rate metrics. Page 11

#### Supplementary Figures

Supplementary Figure 1 | MRC scores for upper limb gestures. Page 14

Supplementary Figure 2 | 3D brain visualization. Page 15

Supplementary Figure 3 | Gesture paradigm. Page 16

Supplementary Figure 4 | Model training and testing sessions. Page 17

Supplementary Figure 5 | Performance scores are calculated in the validation phase. Page 18

Supplementary Figure 6 | Aligning trials as preparation for label assignment. Page 19

Supplementary Figure 7 | Determining label bounds. Page 20

Supplementary Figure 8 | Label assignment for all gestures and Rest. Page 21

Supplementary Figure 9 | Example: $\beta_2$ onset as a proxy for movement onset. Page 22

Supplementary Figure 10 | Classification model architecture. Page 23

Supplementary Figure 11 | Computing modulation amplitude. Page 24

Supplementary Figure 12 | Composite sensitivity, accuracy, and confusion matrices. Page 26

Supplementary Figure 13 | Composite false positive frequencies (FPFs). Page 27

Supplementary Figure 14 | Power trial rasters for Wrist Ext. Page 28

Supplementary Figure 15 | Power trial rasters for ASL-Y. Page 29

Supplementary Figure 16 | Power trial rasters for Splay. Page 30

Supplementary Figure 17 | Power trial rasters for Thumb Flex. Page 31

Supplementary Figure 18 | Power trial rasters for Ring Flex. Page 32

Supplementary Figure 19 | Power trial rasters for Wrist Flex. Page 33

Supplementary Figure 20 | Power trial rasters for Shoulder. Page 34

Supplementary Figure 21 | Power trial rasters for Grasp. Page 35

Supplementary Figure 22 | Change in channel 91's high-gamma power during ASL-Y. Page 36

**Supplementary Tables**

Supplementary Table 1 | Descriptions of attempted gestures. Page 37

Supplementary Table 2 | Offline performance per model. Page 38

Supplementary Table 3 | Summary of offline performance. Page 38

Supplementary Table 4 | Performance metrics per gesture. Page 39

**References** Page 40

### Supplementary Notes

#### Supplementary Note 1. ALS Functional Rating Scale (ALSFRS-R)

The clinical care team at Johns Hopkins Hospital obtained an assessment for the ALSFRS-R[1] measure one week before we started collection of the multi-class gesture data. In total, the study participant in the scored 20 out of 48 points. The scores for each category of the ALSFRS-R are shown below in bold.

**1. SPEECH**

4. Normal speech processes

3. Detectable speech disturbances

2. Intelligible with repeating

**1. Speech combined w/ nonvocal communication**

0. Loss of useful speech

**2. SALIVATION**

4. Normal

3. Slight but definite excess of saliva in mouth; may have nighttime drooling

2. Moderately excessive salvia; may have minimal drooling

**1. Marked excess of saliva with some drooling**

0. Marked drooling; requires constant tissue or handkerchief

**3. SWALLOWING**

4. Normal eating habits

3. Early eating problems – occasional choking

2. Dietary consistency changes

**1. Needs supplemental tube feedings**

0. NPO (exclusively parenteral or enteral feeding)

**4. HANDWRITING**

4. Normal

3. Slow or sloppy; all words are legible

2. Not all words are legible

**1. Able to grip pen but unable to write**

0. Unable to grip pen

**5a. CUTTING FOOD AND HANDLING UTENSILS**

(Patients *without* gastrostomy) **N/A**

4. Normal

3. Somewhat slow and clumsy; no help needed

2. Can cut most foods (clumsy and slow); some help needed

1. Foods must be cut by someone, but can still feed slowly

0. Needs to be fed

**5b. CUTTING FOOD AND HANDLING UTENSILS**

(Patients *with* gastrostomy)

4. Normal

3. Clumsy, but able to perform all manipulations independently

2. Some help needed with closures and fasteners

1. Provides minimal assistance to caregiver

**0. Unable to perform any aspect of task**

**6. DRESSING AND HYGIENE**

4. Normal

3. Independent self-care with effort or decreased efficiency

2. Intermittent assistance or substitute methods

1. Needs attendant for self-care

**0. Total dependence**

**7. TURNING IN BED & ADJUSTING BEDCLOTHES**

4. Normal

3. Somewhat slow and clumsy, but no help needed

2. Can turn alone or adjust sheets, with great difficulty

**1. Can initiate, but not turn or adjust sheets alone**

0. Helpless

**8. WALKING**

**4. Normal**

3. Early ambulation difficulties

2. Walks with assistance

1. Nonambulatory functional movement only

0. No purposeful leg movement

**9. CLIMBING STAIRS**

4. Normal

**3. Slow**

2. Mild unsteadiness or fatigue

1. Needs assistance

0. Cannot do

**10. DYSPNEA** (shortness of breath)

4. None

**3. Occurs when walking**

2. Occurs with one or more of the following: eating, bathing, dressing

1. Occurs at rest, either sitting or lying

0. Significant difficulty, considering using mechanical support

**11. ORTHOPNEA** (trouble breathing when lying flat)

4. None

**3. Some difficulty sleeping at night due to shortness of breath; does not routinely use more than 2 pillows**

2. Needs extra pillows in order to sleep (more than 2)

1. Can only sleep sitting up

0. Unable to sleep

**12. RESPIRATORY INSUFFICIENCY**

4. None

3. Intermittent use of BiPAP/NIV

**2. Continuous use of BiPAP/NIV during the night**

1. Continuous use of BiPAP/NIV during the night and day

0. Mechanical ventilation (intubation or tracheostomy)

**Total: 20/48**

#### Supplementary Note 2. Neural representations of gestures may drift

Neural representations of gestures recorded with ECoG can potentially "drift" (represented by a shift in their location or orientation in a low-dimensional space), though this can result in part due to variations in how attempted gestures are executed across sessions[2]. However, in our study, we had to consider the potential for neural representations to drift due to the progression of ALS. For any set of training sessions, the statistics of the neural representations could have depended on the time span in which the data was collected. Consequently, our class-selection algorithm did not select the same set of control gestures for each set of training sessions. For example, if the neural representation of some gesture $i$ had less overlap with all other gestures (as well as with the Rest class) in sessions 3 and 4 compared to sessions 1 and 2, then gesture $i$'s performance score would have been greater when including sessions 3 and 4 in addition to sessions 1 and 2 during model training and validation.

#### Supplementary Note 3. Label assignment using the 70% threshold crossing

We chose to apply the gesture labels according to a percentage threshold value and not, for example, according to the visual onset and offset of neural activity. This was done to avoid applying gesture labels to samples close to the onset and offset times because by visual inspection, features close to these time points were not sufficiently different from Rest. Such labeling may have produced a large class overlap between the gesture classes and Rest, which may have subsequently reduced the discriminatory power of the classifier. Therefore, we chose to label the data according to a threshold value in order to minimize this class overlap. We chose the 70% value heuristically, based on results from preliminary analysis. After all gesture labels were applied, the remaining samples were labeled as Rest.

#### Supplementary Note 4. Min() and max() functions for performance score

If gestures $i$ and $j$ had FPFs of 0.01 $hr^{-1}$ and 1 $hr^{-1}$, respectively (with equal sensitivity, accuracy, and precision), gesture $i$'s performance score would have been 100 times larger than gesture $j$'s. This would have unfairly biased gesture $i$ to be selected as a control gesture over gesture $j$. As such, we applied ceilings of 90%, 80%, and 80% (using the $\min(\ )$ function) for sensitivity, accuracy, and precision, respectively, and a floor of 1 $hr^{-1}$ (using the $\max(\ )$ function) for FPF. The ceiling and floor values were selected qualitatively as criteria for good BCI performance and based on the performance targets we specified in our clinical trial. We did not include the latency in the calculation of our performance score because we aimed to primarily target sensitivity, accuracy, and FPF, and because we observed less variance in per-gesture latency in our preliminary analyses.

### Supplementary Methods

#### Supplementary Method 1. Clinical trial inclusion and exclusion criteria

| Inclusion Criteria |
| --- |
| • Complete or incomplete tetraplegia (quadriplegia), tetraparesis (quadriparesis), severe ataxia, or moderate to severe motor impairments in both upper limbs, based on neurological exam. In addition, these motor impairments may be combined with severe motor-related speech impairment (dysarthria or anarthria), as in Locked In Syndrome (LIS) and amyotrophic lateral sclerosis (ALS), including the bulbar variant of ALS.<br>• Clinical diagnosis established for the etiology of motor impairments, including brainstem stroke1, spinal cord injury, or progressive and irreversible neuromuscular disease, including amyotrophic lateral sclerosis (ALS).<br>• Persistence of motor impairments at least one year prior to enrollment if due to stroke or spinal cord injury<br>• 22-70 years<br>• Meeting surgical safety criteria, including surgical clearance by the participant's primary healthcare provider, study physicians, and any necessary consultants<br>• Ability to communicate reliably, such as through eye movement<br>• Willingness and ability to provide informed consent<br>• Screened by rehabilitation psychologist with a result showing that the participant has a stable psychosocial support system with caregiver capable of monitoring participant throughout the study<br>• Ability and willingness to travel up to 100 miles to study location up to three days per week for the duration of the study<br>• Ability to understand and comply with study session instructions<br>• Participant consents to the study and still wishes to participate at the time of the study |
| Exclusion Criteria |
| • Performance on formal neuropsychological testing that indicates a significant psychiatric disorder or cognitive impairments that would interfere with obtaining informed consent or fully participating in study activities.<br>• Suicide attempt or persistent suicidal ideation within the past 12 months.<br>• Implanted devices that are incompatible with MRI, which may include pacemakers, cardiac defibrillators, spinal cord or vagal nerve stimulators, deep brain stimulators, and cochlear implants.<br>• History of substance abuse, narcotic dependence, or alcohol dependence in past 24 months<br>• Medical conditions contraindicating surgery of a chronically implanted device (e.g. osteomyelitis, diabetes, hepatitis, any autoimmune disease/disorder, epilepsy, skin disorders |

causing excessive skin sloughing or poor wound healing, blood or cardiac disorder requiring chronic anti-coagulation)

• Other chronic, unstable medical conditions that could interfere with subject participation.

• Presence of pre-surgical findings in anatomical, functional, and/or vascular neuroimaging that makes achieving implant locations within desired risk levels too challenging (to be decided by neurological and neurosurgical team)

• Prior cranioplasty

• Inability to undergo MRI or anticipated need for an MRI during the study period

• Participants with active infections or unexplained fever

• Participants with other morbid conditions making the implantation of the recording elements unsafe; not limited to: significant pulmonary, cardiovascular, metabolic, or renal impairments making the surgical procedure unsafe

• Pregnancy (confirmation through blood test)

• Nursing an infant, planning to become pregnant, or not using adequate birth control

• Corrected vision poorer than 20/100

• HIV or AIDS infection

• Existing scalp lesions or skin breakdown

• Chronic oral or intravenous use of steroids or immunosuppressive therapy

• Active cancer within the past year or requires chemotherapy

• Uncontrolled autonomic dysreflexia within the past 3 months

• Hydrocephalus with or without an implanted ventricular shunt

• Participants in whom it is medically contraindicated to stop anti-coagulant medications during surgery

#### Supplementary Method 2. Selecting control gestures for model testing

We performed leave-one-session-out cross-validation on each set of training data, $D_{train}$, where each validation fold, $D_{valid\ fold}$, consisted of one session from $D_{train}$ while the folds for training the corresponding cross-validation model, $D_{train\ folds}$, consisted of all other sessions (Supplementary Fig. 5a). Further, $D_{train\ folds}$ and $D_{valid\ fold}$ were balanced to contain an equal number of samples per class by down-sampling all classes to match the class with the fewest samples. Each session in $D_{train}$ was used once as a validation fold.

For each validation fold, $D_{valid\ fold}$, we generated the output probabilities for the Rest class and for all 13 gesture classes (see *Model architecture and training*) using the corresponding 14-class cross-validation model (Supplementary Fig. 5b). Output probabilities were computed in 100 ms increments (see *Spectral power computation*) and smoothed by taking the average of the most recent 1 s. When the smoothed Rest probability dropped below 0.05 (chosen heuristically) the gesture with the highest probability was classified (Supplementary Fig. 5b). We evaluated sensitivity, accuracy, precision, and false positive frequency (FPF) for each gesture (Supplementary Method 3), which together composed a performance score (main text, eq. 1) for that gesture across all validation folds.

For each $D_{train}$, the six gestures with the highest performance scores were designated as control gestures (i.e., gestures that could have been used to control an assistive application using BCI) (Supplementary Fig. 5c). We chose to use six gestures in order to simulate directional control commands (such as "up," "down," "left," "right," "enter," and "back"). The control gestures were used to train a model for simulating online testing. The other seven gestures were designated as "excluded gestures," and were excluded from training the model for simulating online testing. A set of six control gestures and seven excluded gestures were determined for each set of training data, $D_{train}$.

#### Supplementary Method 3. Performance metrics

Sensitivity for any control gesture $i$ was measured as the percentage of movement attempts for gesture $i$ that generated true positive detections of any gesture (i.e., including, but not limited to, gesture $i$). For a detection to be a true positive, it must have occurred within a detection window of 2.5 s that started at the onset of $\beta_2$ modulation ($t_{gesture\ \beta 2\ onset}$) for any given trial where 2.5 s was sufficient time to complete an attempted gesture. Trials with no detections during the detection window were considered false negatives. The sensitivity is defined in eqs. S1 and S2.

$$Sens_i = \frac{N_{TP\ |\ i\ attempt}}{N_{i\ attempt}} \times 100\% \qquad \text{(eq. S1)}$$

$$Sens_i = \frac{N_{TP\ i\ |\ i\ attempt} + N_{TP\ \sim i\ |\ i\ attempt}}{N_{i\ attempt}} \times 100\% \qquad \text{(eq. S2)}$$

where $N_{TP\ |\ i\ attempt}$ was the number of true positive classifications (of any gesture) that occurred during gesture $i$ attempts (given gesture $i$ attempts). $N_{i\ attempt}$ was the total number of gesture $i$ attempts and $N_{i\ attempt} \geq N_{TP\ |\ i\ attempt}$. Moreover, $N_{TP\ |\ i\ attempt}$ was composed of $N_{TP\ i\ |\ i\ attempt}$ and $N_{TP\ \sim i\ |\ i\ attempt}$, which were the number of true positive classifications of gesture $i$ during a gesture $i$ attempt, and true positive classifications of other gestures (not gesture $i$) during a gesture $i$ attempt, respectively. A classification for any gesture $i$ that occurred outside of the detection window was considered a false positive.

The false positive frequency (FPF) for gesture $i$ was defined as the number of false positive gesture $i$ classifications per unit time. Note that it was not possible to compute the specificity and false positive rate (FPR) as percentage values because these metrics both depend on a quantity of true negative classifications, which could not be defined in the continuous time domain (see Supplementary Method 5). We defined the FPF in eq S3.

$$FPF_i = \frac{N_{FP\ i}}{T} \qquad \text{(eq. S3)}$$

where $N_{FP\ i}$ is the number of gesture $i$ false positives and $T$ was the recording time in which the FPF was measured. During cross-validation, $T$ was the total time of all 5 blocks in a particular validation fold (a validation fold consisted of a single session during which 5 blocks were recorded). However, when we performed model testing, we subtracted the detection window for every trial in which an excluded gesture was attempted. This was to account for any classifications generated as a result of an excluded gesture attempt (because the testing sessions were structured in the same format as the training sessions, they contained all visual gesture cues and corresponding gesture attempts by the participant, including the excluded gestures). This adjustment primarily affected the FPF computation as the recording time $T$ was reduced by 100 s per block (10 excluded gestures x 4 trials/excluded gestures x 2.5 s/trial).

The accuracy for any control gesture $i$ was the percentage of gesture $i$ attempts that were correctly classified as gesture $i$. Specifically, accuracy was measured as a ratio of true positive gesture $i$ classifications that occurred during gesture $i$ attempts ($N_{TP\ i\ |\ i\ attempt}$) to all true positive classifications that occurred during gesture $i$ attempts ($N_{TP\ |\ i\ attempt}$). The accuracy is defined in eqs. S4 and S5.

$$Acc_i = \frac{N_{TP\ i\ |\ i\ attempt}}{N_{TP\ |\ i\ attempt}} \qquad \text{(eq. S4)}$$

$$Acc_i = \frac{N_{TP\ i\ |\ i\ attempt}}{N_{TP\ i\ |\ i\ attempt} + N_{TP\ \sim i\ |\ i\ attempt}} \qquad \text{(eq. S5)}$$

The precision for any control gesture $i$ was the percentage of gesture $i$ classifications that were caused by gesture $i$ attempts. Specifically, precision was measured as a ratio of true positive gesture $i$ classifications that occurred during gesture $i$ attempts to all gesture $i$ true positives (occurring during gesture $i$ attempts or during attempts of another gesture). The precision is defined in eqs. S6 and S7.

$$Prec_i = \frac{N_{TP\ i\ |\ i\ attempt}}{N_{TP\ i}} \qquad \text{(eq. S6)}$$

$$Prec_i = \frac{N_{TP\ i\ |\ i\ attempt}}{N_{TP\ i\ |\ i\ attempt} + N_{TP\ i\ |\ \sim i\ attempt}} \qquad \text{(eq. S7)}$$

where $N_{TP\ i}$ is the number of all gesture $i$ true positives regardless of whether they occurred during an attempt of gesture $i$ ($N_{TP\ i\ |\ i\ attempt}$) or any other gesture ($N_{TP\ i\ |\ \sim i\ attempt}$). Note that a decrease in the precision for gesture $i$ can also cause a decrease in the accuracy for some other gesture $j$ due to the same underlying classification errors. This is because an increase in $N_{TP\ i\ |\ \sim i\ attempt}$ (misclassification term in the denominator of eq. S7) where $\sim i = j$ will lower the precision for gesture $i$ and while simultaneously lowering the accuracy for gesture $j$ due to an increase in the $N_{TP\ \sim j\ |\ j\ attempt}$ term (misclassification term in the denominator of eq. S8) where $\sim j = i$. Therefore, when accounting for misclassifications between all pairs of true and predicted classes, this produces a positive correlation between overall model accuracy and overall (macro-average precision).

Latency for each classification was defined as the time delay from onset of $\beta_2$ modulation to gesture classification. For the n[th] true positive classification following the k[th] trial of an attempted control gesture $i$, the latency is defined as

$$lat_i^{(n)} = t_{classification}^{(n)} - t_{gesture\ i\ \beta 2\ onset}^{(k)} \qquad \text{(eq. S8)}$$

For example, the first true positive classification (n=1) could have occurred during the second trial (k=2) of gesture $i$. The mean latency for gesture $i$ is the mean over all classifications for gesture $i$ attempts.

$$Lat_i = \frac{1}{N_{TP\,|\,i\,attempt}} \sum_n^{N_{TP\,|\,i\,attempt}} lat_i^{(n)} \qquad \text{(eq. S9)}$$

where $lat_i^n$ is the n[th] true positive classification generated by a gesture $i$ attempt.

#### Supplementary Method 4. Lack of specificity and false positive rate metrics

It was not possible to compute the specificity (true negative rate, TNR) and the false positive rate (FPR) because both depend on quantifying the number of true negative classifications. Briefly, specificity and FPR are defined below as percentage values:

$$TNR = \frac{TN}{FP+TN} = 1 - FPR \qquad \text{(eq. S10)}$$

$$FPR = \frac{FP}{FP+TN} \qquad \text{(eq. S11)}$$

where $FP$ and $TN$ are the number of false positives and true negatives, respectively. Though we were able to count the number of FPs as defined in Supplementary Method 3, the concept of a TN (identifying the instance when a classification is correctly omitted) is ill-posed in the continuous time domain because there do not exist discrete (instantaneous, not continuous) countable events of correctly omitted classifications.

### Supplementary Figures

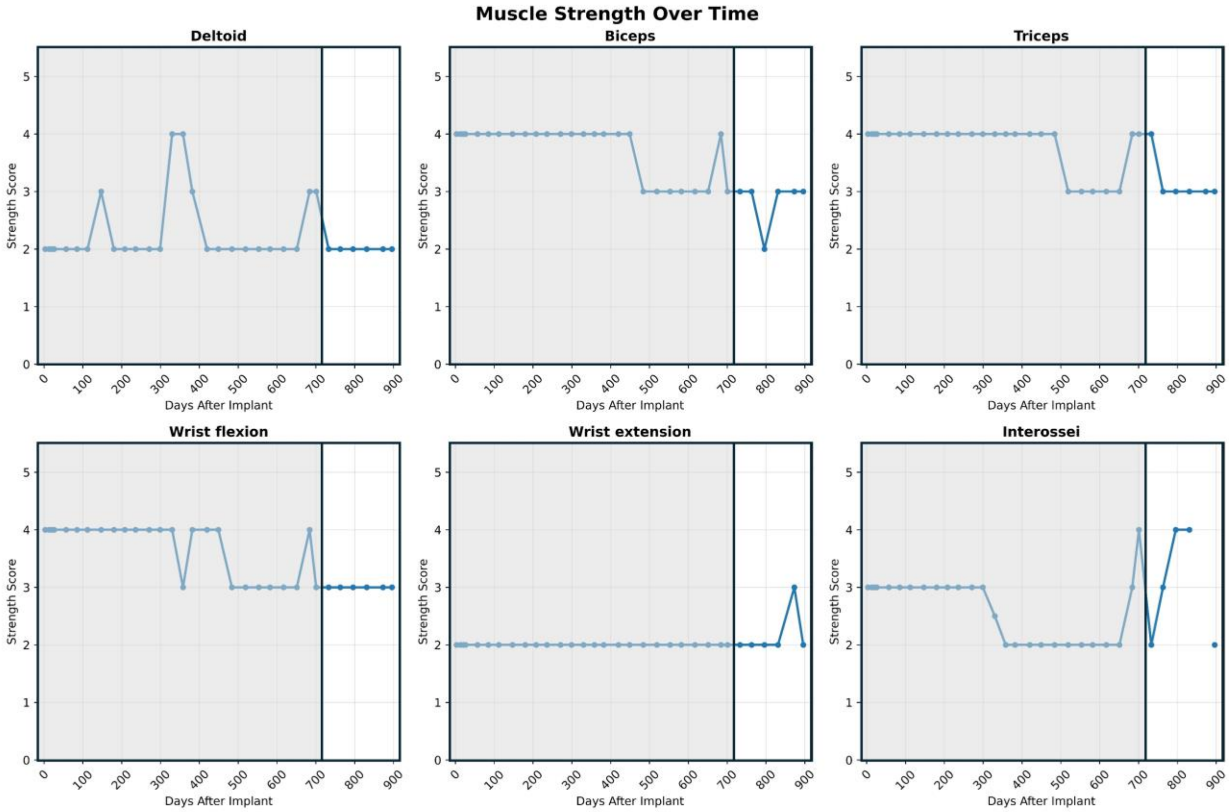


**Supplementary Figure 1 | MRC scores for upper limb muscles.** All scores for the upper limb muscles that were recorded for our study participant during the clinical trial are shown relative to the first day of implant. The period in highlighted period in white represents the 5-month period of this study.

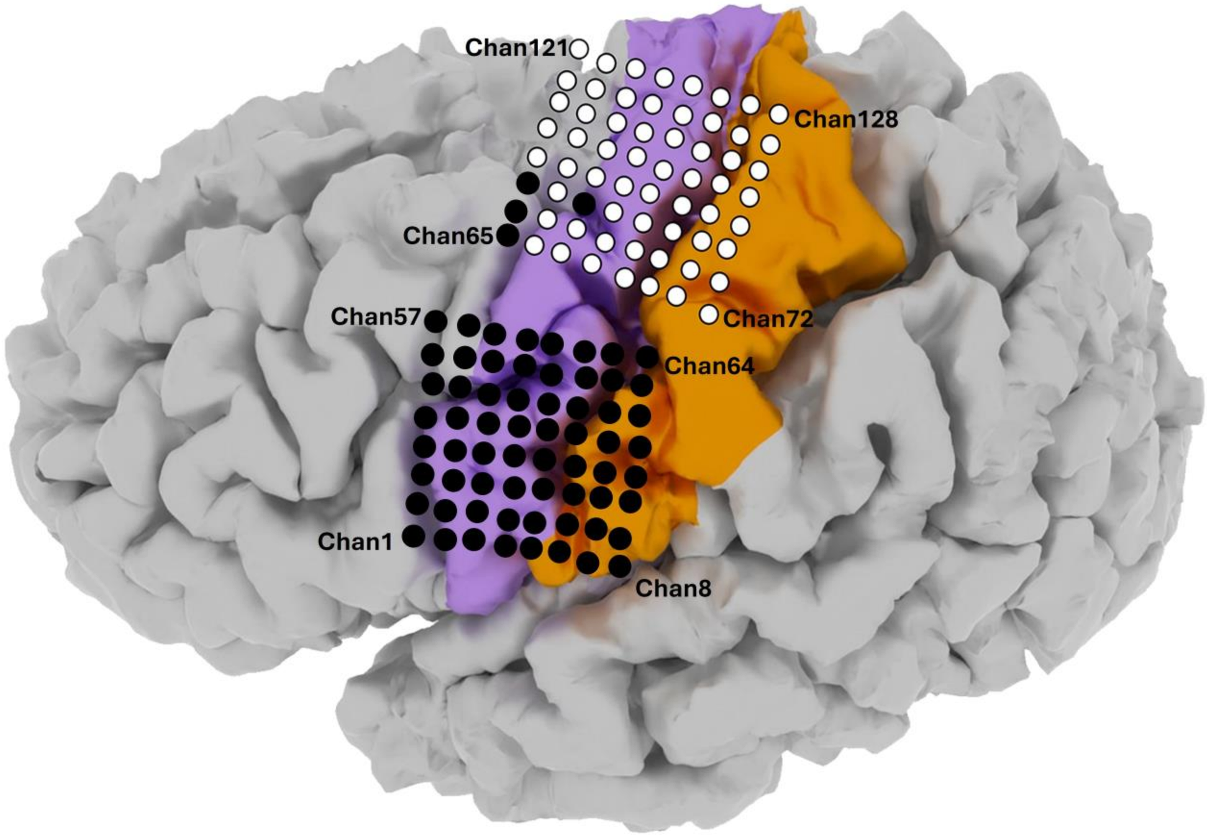


**Supplementary Figure 2 | 3D brain visualization.** Visualization of both 64-electrode grids on a 3D reconstruction of the participant's left cortical surface. The dorsal and ventral grids primarily covered the cortical upper limb and face regions, respectively. Electrodes were sequentially numbered from left to right and bottom to top. The black electrodes were excluded from analysis. Color coding of the cortex indicates anatomical landmarks: magenta for pre-central gyrus and orange for post-central gyrus.

a
b
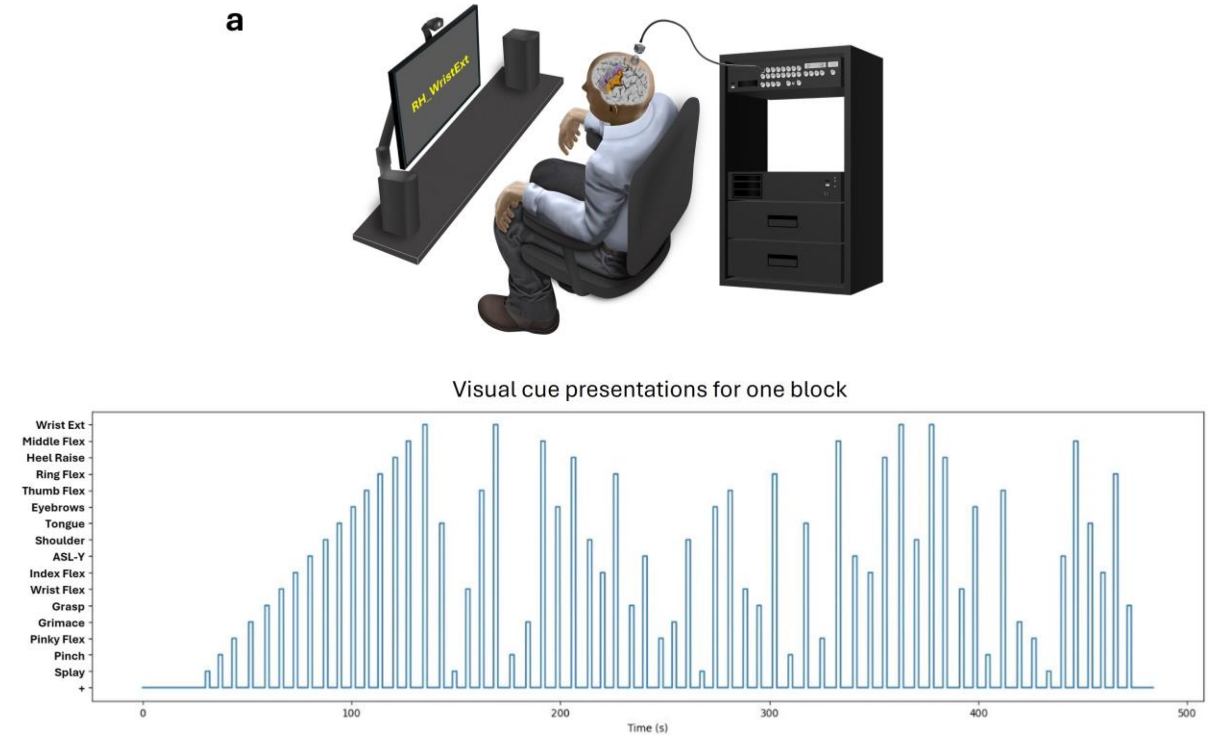


**Supplementary Figure 3 | Gesture paradigm.** (**a**) As the participant was seated in a chair with his forearms on the armrests and his feet planted on the floor, he was instructed to attempt a brisk facial, right leg, or right upper limb gesture immediately after the visual stimulus describing that gesture appeared on the monitor. One trial consisted of one 2 s visual text stimulus followed by an interstimulus interval (ISI) during which a white crosshair (+) appeared in the center of the monitor. The length of each ISI was randomly chosen from a uniform distribution bounded between 4 - 6 s to reduce anticipatory behavior. In total each trial lasted between 6 - 8 s. (**b**) An example of the pseudo-random interleaved visual cue presentations for one block where the blue lines represent when the visual cues were presented.

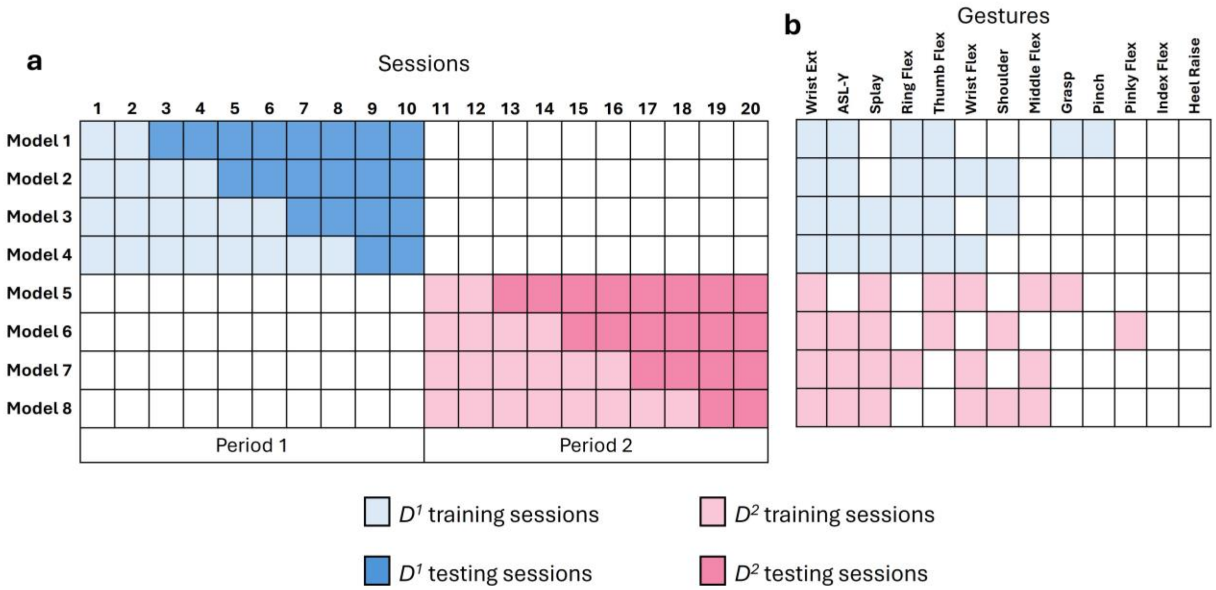


**Supplementary Figure 4. Model training and testing sessions.** (**a**) Each of the 8 rows in the table corresponds to a model trained on a particular dataset, $D_{train}$ (a subset of $D^1$ or $D^2$) and tested on a different dataset, $D_{test}$. For example, Model 2 was trained on sessions 1-4 (light blue cells) was tested on sessions 5-10 (dark blue cells). Similarly, Model 8 was trained on sessions 11-18 (light pink cells) and was tested on sessions 19-20 (dark pink cells). Sessions 1-10 and 11-20 were recorded in Periods 1 and 2, respectively. (**b**) For each set of training sessions in the rows of (**a**), the six control gestures used for model training are highlighted. Index Flex and Heel Raise were never selected as control gestures for any model, though this was because of their poor performance in the validation phase (see Supplementary Method 3) and not because they were manually excluded.

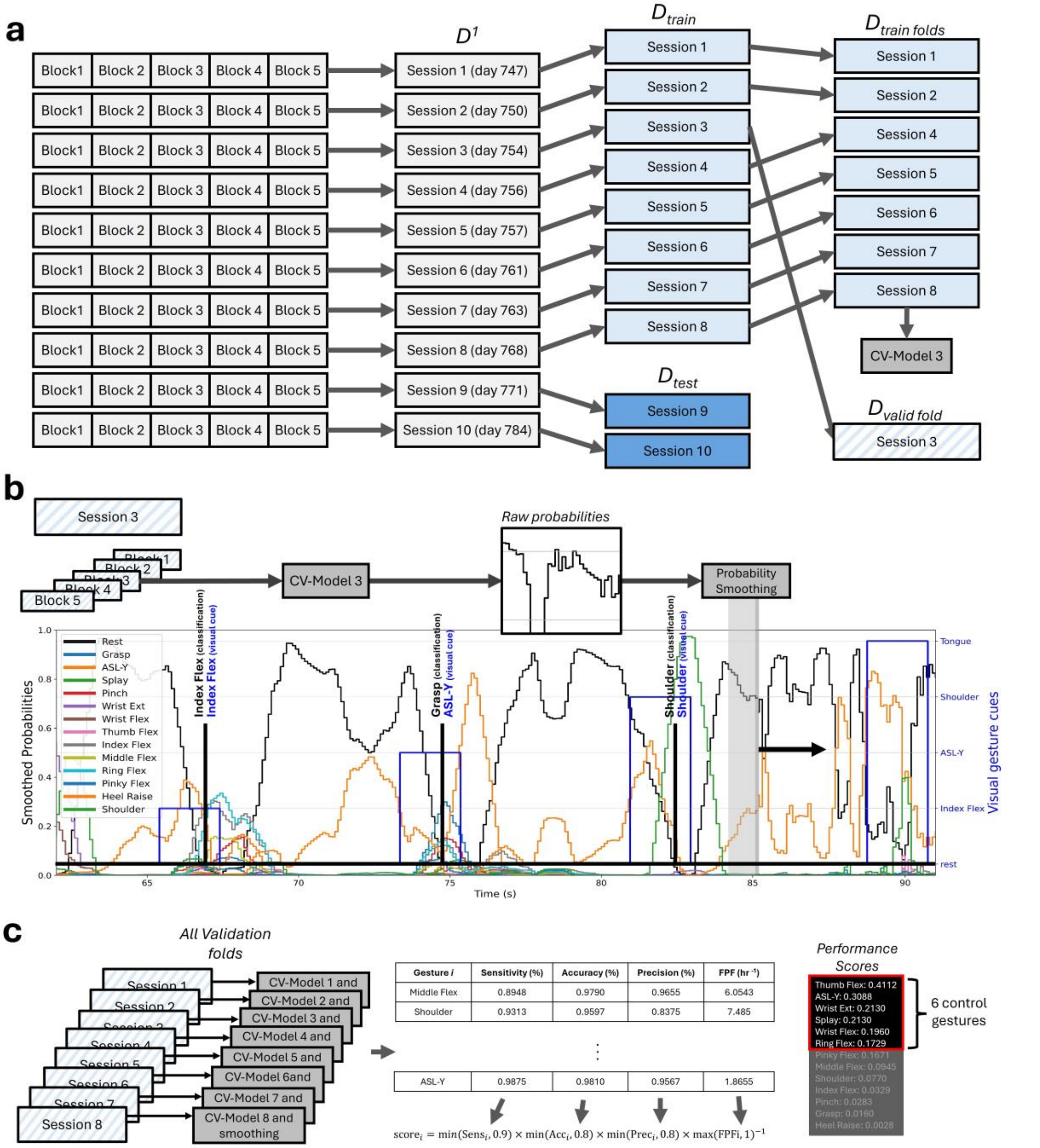


**Supplementary Figure 5 | Performance scores are calculated in the validation phase.** As an example, all sessions from Period 1 are used for illustrating cross-validation and subsequent calculation of the performance scores (**a**) For Period 1, all sessions were collectively denoted as $D^1$, which was partitioned into $D_{train}$ and $D_{test}$. In this example, $D_{train}$ consists of sessions 1 - 8 and $D_{test}$ consists of sessions 9 and 10. $D_{train}$ is then partitioned into $D_{train\ folds}$ (in this illustration, sessions 1,2, 4-8) and $D_{valid\ fold}$ (session 3). Cross-validation (CV)-Model 3, corresponding to validation session 3, was created from $D_{train\ folds}$. (**b**) All 5 blocks of the validation data are evaluated by CV-Model 3, from which the probabilities of all gestures were computed and smoothed by taking the average of the most recent 1 s (most recent 10 output probabilities for each class). The vertical black lines denote when the Rest class fell below the 0.05 threshold, and the class with the highest smoothed probability was selected. The blue lines represent the presentation of the visual cues. (**c**) For each validation fold, probabilities were output by their corresponding CV-Models, smoothed, and used to generate classifications. Then the overall performance metrics for each gesture are computed. Using these metrics the performance score is then calculated for each gesture and the six gestures with the highest performance scores are selected as control gestures.

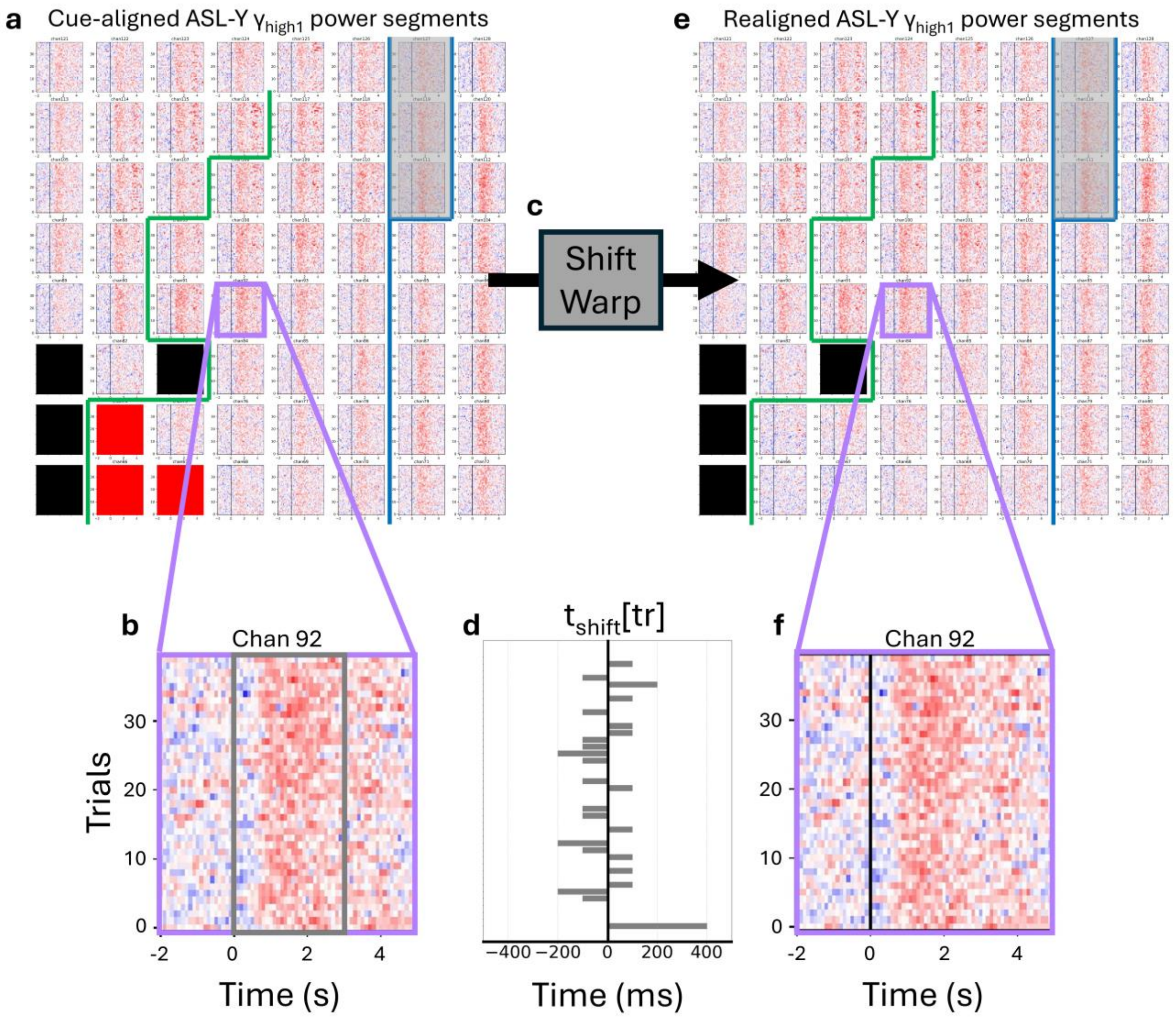


**Supplementary Figure 6 | Realigning trials as preparation for label assignment.** In order to train models for validation and testing, we assigned labels to each sample of the training data. We concatenated segments of $\gamma_{high1}$ power across trials (-2 to 5 s relative to visual cue onset), resulting in $\gamma_{high1}$ power segments that were aligned to the onset of the visual cue (cue-aligned). However, to account for the inter-trial variability of response latencies across trials, the $\gamma_{high1}$ power segments were realigned using a shift warping model. This realignment was performed as the first step in the sample labeling process. As an example, this figure illustrates segment realignment for only ASL-Y trials. Specifically, these trials were part of the set of training data consisting of sessions 1 and 2, each with 20 ASL-Y trials. (**a**) Segments of cue-aligned $\gamma_{high1}$ power are concatenated across all 40 ASL-Y trials for all channels with significant peak modulation specific to ASL-Y. Channels without significant peak modulation are marked in bright red and those excluded from analysis are marked in black. Channels are numbered sequentially from 65 to 128, as shown Supplementary Fig. 2. The approximate central sulcus location is delineated by a thick blue line (CS) and widens at the top such that channels 111, 119, and 127 are over it. The pre-central sulcus is delineated by a thick green line (Pre-CS). (**b**) Channel 92 is magnified to better illustrate the cue-aligned segments of $\gamma_{high1}$ power. Power segments range from -2 to 5 s relative to the onset of the visual gesture cue (at 0 s). For each channel with significant peak modulation, the cue-aligned trial subsegments from 0 to 3 s (within the grey box) were used to train a shift-warping model in (**c**). (**c**) The shift warping model was used to realign $\gamma_{high1}$ power segments by appropriately shifting each trial by a positive or negative trial-specific time shown in (**d**). Note that no temporal warping was performed. (**e**) For all channels (including those without significant peak modulation), each segment of $\gamma_{high1}$ power was shifted by its trial-specific time. (**f**) Channel 92 is magnified to better illustrate the shifted segments of $\gamma_{high1}$ power. For example, the 400 ms shift for the first (bottom) trial segment is most noticeable in this example.

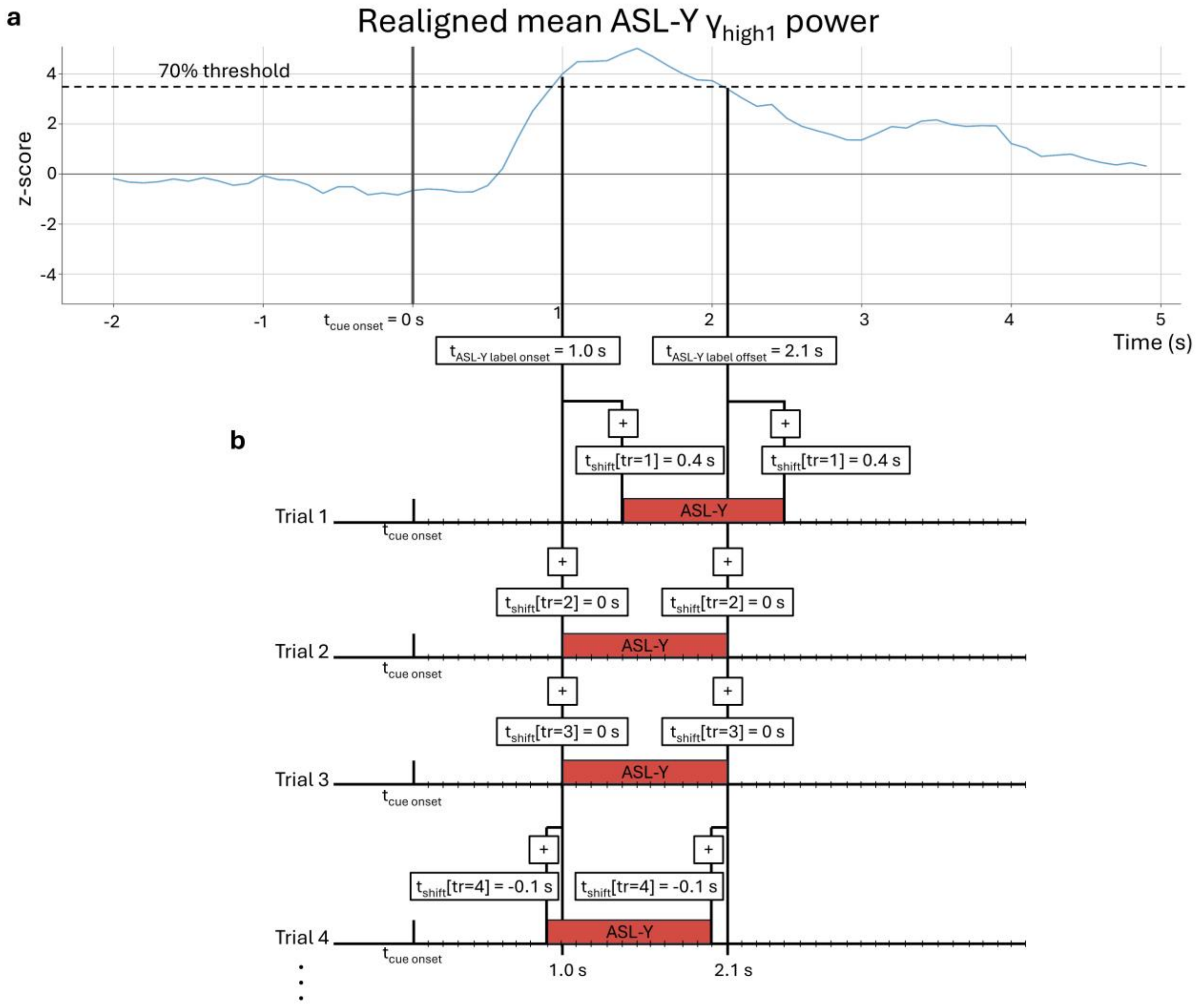


**Supplementary Figure 7 | Determining label bounds.** In order to train models for validation and testing, we assigned labels to each sample of the training data. Following realignment of $\gamma_{high1}$ power segments, the mean $\gamma_{high1}$ power was used to determine the labeling bounds. As an example, this process is illustrated for the ASL-Y trials which were previously described in Supplementary Fig. 6. (**a**) After alignment, the mean $\gamma_{high1}$ power was computed across all channels with significant peak modulation specific to ASL-Y. Then the two closest time points to where the mean $\gamma_{high1}$ power crossed above and below the threshold z-score value (70% of the maximum of the mean $\gamma_{high1}$ power) were determined. In this example, these two time points were $t_{ASL\text{-}Y\ label\ onset}$ = 1.0 s and $t_{ASL\text{-}Y\ label\ offset}$ = 2.1 s, respectively, relative to cue onset. (**b**) We then determined the labeling bounds for each ASL-Y trial (i.e., the time points between which all the trial samples would be labeled as ASL-Y) and accounted for the trial-specific shift described in Supplementary Fig. 6. Specifically, for each trial, the labeling bounds were $t_{ASL\text{-}Y\ label\ onset} + t_{shift}[tr]$ and $t_{ASL\text{-}Y\ label\ offset} + t_{shift}[tr]$. For example, for ASL-Y trial 4, all samples between and including 0.9 s and 2.0 s relative to onset of the gesture cue were labeled as ASL-Y.

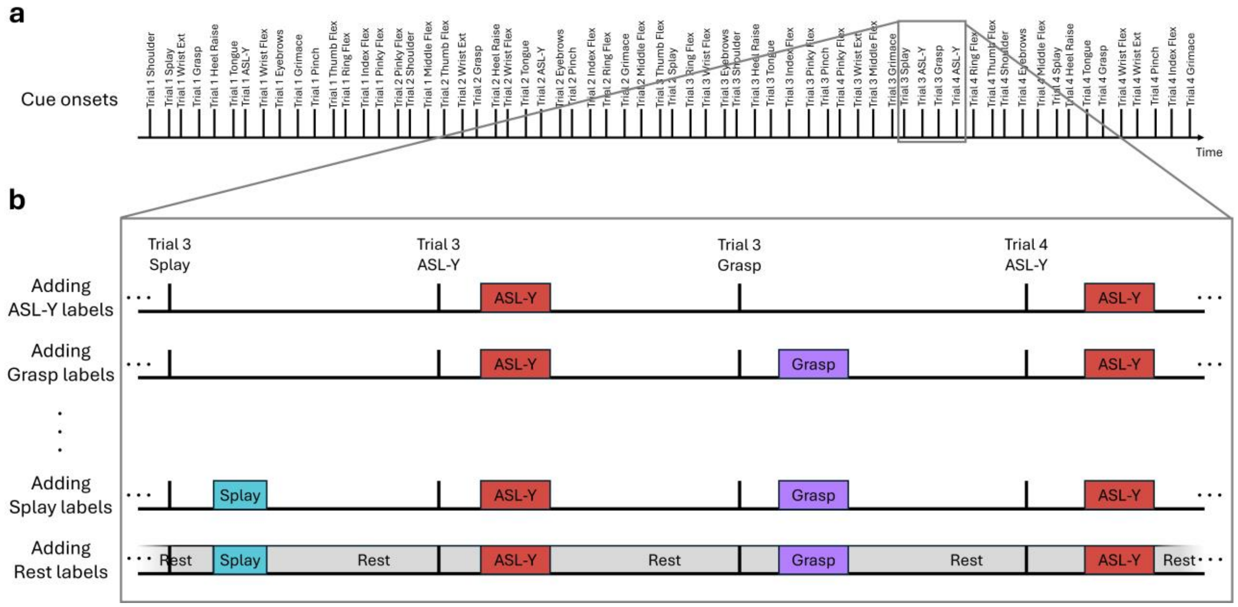


**Supplementary Figure 8 | Label assignment for all gestures and Rest.** In order to train models for validation and testing, we assigned labels to each sample of the training data. The processes illustrated in Supplementary Figs. 6 and 7 were first repeated for each gesture. This figure illustrates the label assignment for one block (of one session) of the example training dataset that was previously described in Supplementary Fig. 6 (sessions 1 and 2). The onsets of the cue gestures are shown in (**a**). One block of training data consisted of 4 trials per gesture (64 trials in total) and was roughly 8 min in duration. (**b**) The samples for each trial were labeled according to each gesture's label onset and offset time points as well as the trial-specific time shift illustrated in Supplementary Figs. 7a and b. After all gesture labels were applied, the remaining samples were labeled as Rest. All blocks in the training data were labeled in this manner.

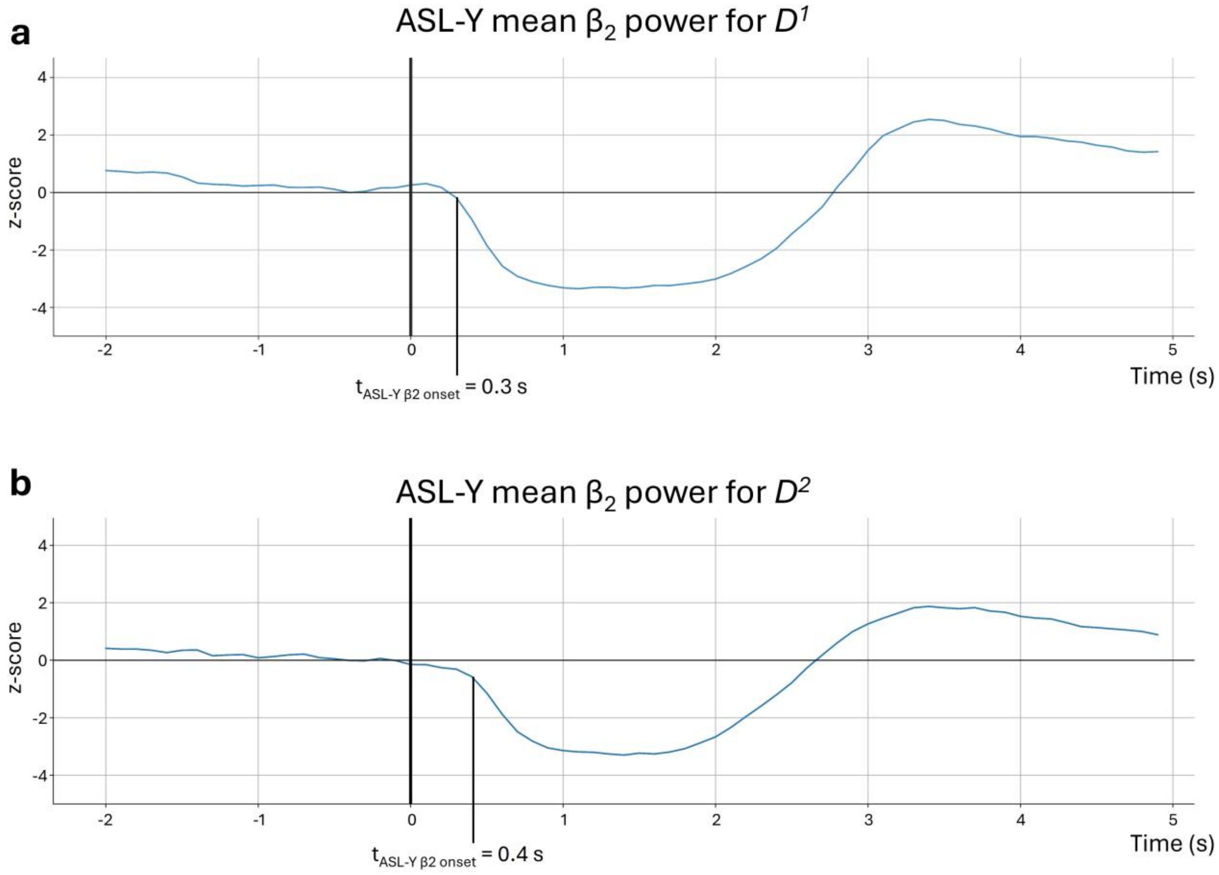


**c**

$\beta_2$ onset times

| | Wrist Ext | ASL-Y | Splay | Ring Flex | Thumb Flex | Wrist Flex | Shoulder | Middle Flex | Grasp | Pinch |
|---|---|---|---|---|---|---|---|---|---|---|
| Period 1 | 0.1 | 0.3 | 0.5 | 0.3 | 0.3 | 0.1 | 0.2 | Not used | 0.4 | 0.3 |
| Period 2 | 0.2 | 0.4 | 0.4 | 0.2 | 0.2 | 0.1 | 0.3 | 0.3 | 0.3 | Not used |

**Supplementary Figure 9 | Example: $\beta_2$ onset as a proxy for movement onset**. We repeated the following process for each gesture (in this figure, ASL-Y is used as an example). First, for each channel, we concatenated segments of cue-aligned $\beta_2$ power (-2 to 5 s relative to the onset of the visual cue) from all 200 trials in Period 1 and all 200 trials in Period 2, respectively. Then, to account for variability in response delay, we realigned them using two shift-warping models[3] (one for each period), trained on channels with significant $\beta_2$ modulation. We then computed the averaged realigned $\beta_2$ power (z-scored) for Period 1 (**a**) and for Period 2 (**b**). The vertical black line represents the onset of the visual cue, which lasted 2 s. For Period 1 and Period 2, the modulation onset times were $t_{ASL\text{-}Y\ \beta 2\ onset}$ = 0.3 s and $t_{ASL\text{-}Y\ \beta 2\ onset}$ = 0.4 s, respectively. Because re-alignment was performed, the ASL-Y modulation onset times for each trial were 0.3 s+ $t_{shift}$[tr] and 0.4 + $t_{shift}$[tr] for Periods 1 and 2 respectively. This method was applied to compute the modulation onset times for all gestures in each period. (**c**) $\beta_2$ onset times relative to the onset of the visual cue are shown for each control gesture that was selected at least once across both Periods 1 and 2. $\beta_2$ onset times for gestures that were never selected as control gestures in a specific period are labeled as "Not used" and $\beta_2$ onset times are not shown for gestures that were never selected as control gestures in either period.

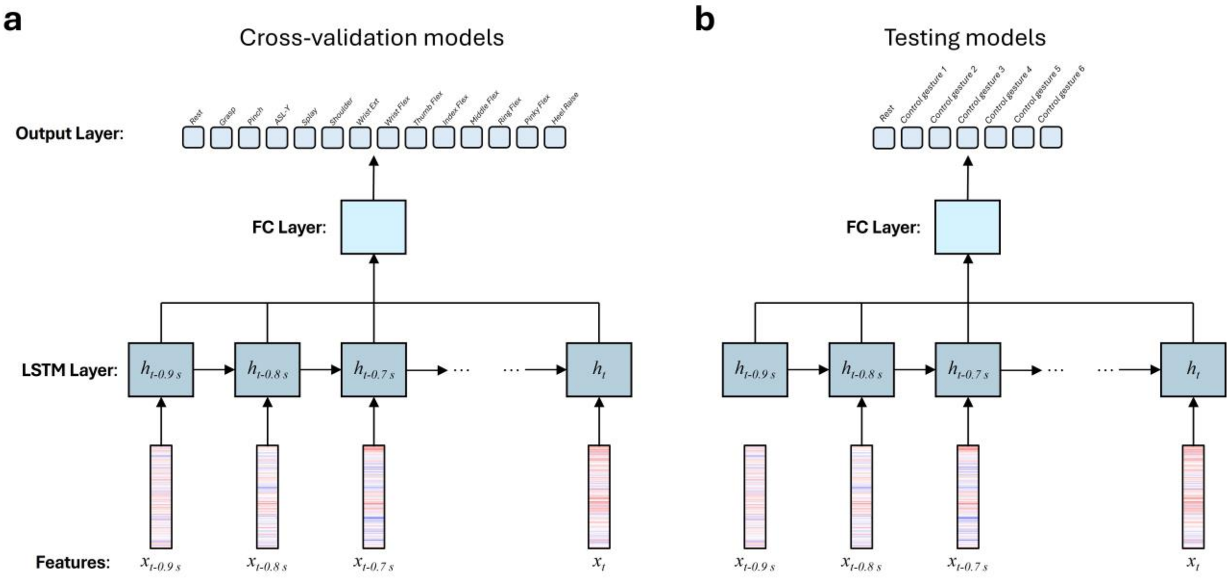


**Supplementary Figure 10 | Classification model architecture.** We implemented a many-to-one recurrent neural network with long short-term memory (LSTM) cells to predict the output probability for each gesture class and Rest. The network architecture consisted of three layers: an LSTM layer with 25 units, followed by a fully-connected layers (10 units) using eLU activation functions for non-linear transformations. Based on whether the model was trained with all 13 gestures and Rest (for cross-validation) or with the 6 control gestures and Rest (for model testing) the output passed through a final fully connected layer with 14 units (**a**) or 7 units (**b**), respectively. A softmax activation function was used for non-linear transformations. Training was performed on 1 s band power sequences with 100 ms frames using backpropagation through time (BPTT). We employed the Adam optimizer with a 10e-4 initial learning rate, training for 75 epochs with batches of 45 samples.

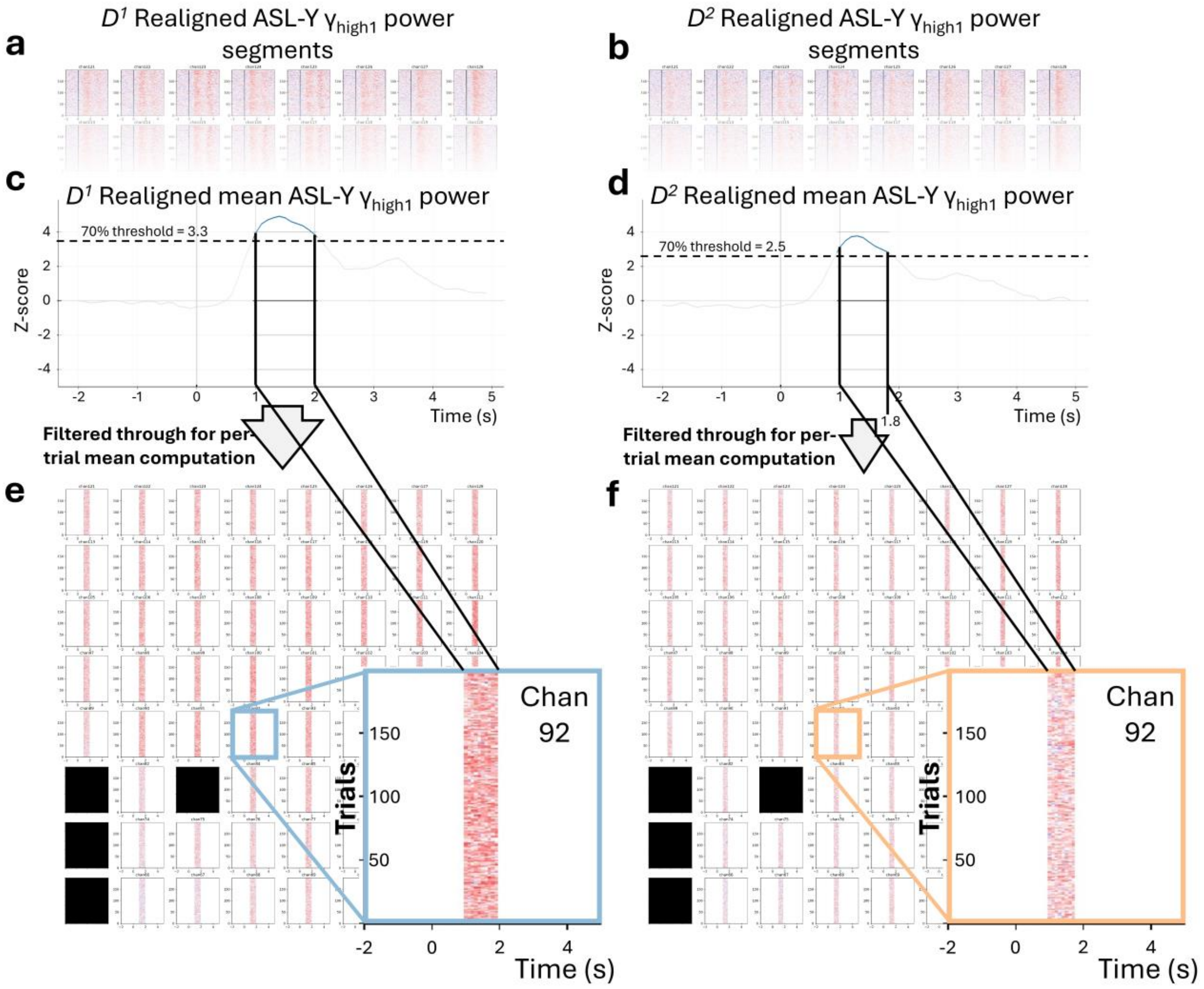


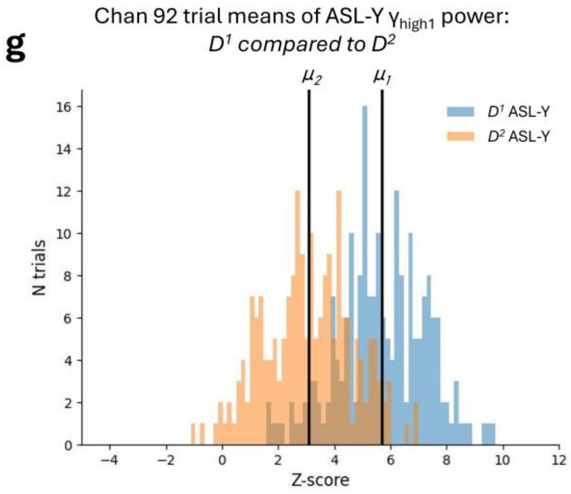


**h** Per-gesture time bounds of 70% threshold crossings

| | Period 1 | Period 2 |
|---|---|---|
| Wrist Ext | [1.4, 1.9] | [1.4, 1.9] |
| ASL-Y | [1.0, 2.0] | [1.0, 1.8] |
| Splay | [1.1, 1.6] | [1.1, 1.6] |
| Thumb Flex | [0.8, 1.5] | [0.9, 1.5] |
| Ring Flex | [1.0, 1.8] | [1.0, 1.8] |
| Wrist Flex | [1.5, 1.8] | [1.5, 1.8] |
| Shoulder | [1.1, 1.4] | [1.1, 1.4] |
| Grasp | [1.0, 1.4] | [1.0, 1.4] |

**Supplementary Figure 11 | Computing modulation amplitude.** We repeated the following process for each gesture (in this figure, ASL-Y is used as an example). First, for each channel, we concatenated segments of cue-aligned $\gamma_{high1}$ power (-2 to 5 s relative to the onset of the visual cue) from all 200 trials in Period 1 and all 200 trials in Period 2. Then, to account for variability in response delay, we realigned the trials using two shift-warping models[3] (one for each period), trained on channels with significant $\gamma_{high1}$ modulation. The realigned trials are shown in (**a**) and (**b**), respectively. We chose to perform trial realignment using only $\gamma_{high1}$-based shift-warping models for consistency with our model training and labeling processes. (**c**, **d**) For both periods, we then computed the mean $\gamma_{high1}$ power across all realigned trials and all channels with significant $\gamma_{high1}$ modulation. We then determined the two time points relative to the onset of the visual cue where this mean $\gamma_{high1}$ power crossed the 70% threshold of the peak value (see *Frequency band selection*). After determining the 70% threshold value, we used the signals between the two time points at the threshold crossing (shown in **e** and **f**) to compute each trial's mean activity for each channel (channels 65, 73, 81, and 83 marked in black were excluded from analysis). For Periods 1 and 2, the insets show per-trial $\gamma_{high1}$ activity for channel 92 that occurred only above the 70% threshold (between 1.0 - 2.0 s, and 1.0 - 1.8 s for Periods 1 and 2, respectively). (**g**) The distributions of trial means for channel 92 during Periods 1 and 2 are shown as an example. A Welch's t-test was performed to compare if the means of these distributions were significantly different (see main text, Fig. 4). (**h**) For each gesture, the time points where the mean $\gamma_{high1}$ power crosses the 70% threshold are shown.

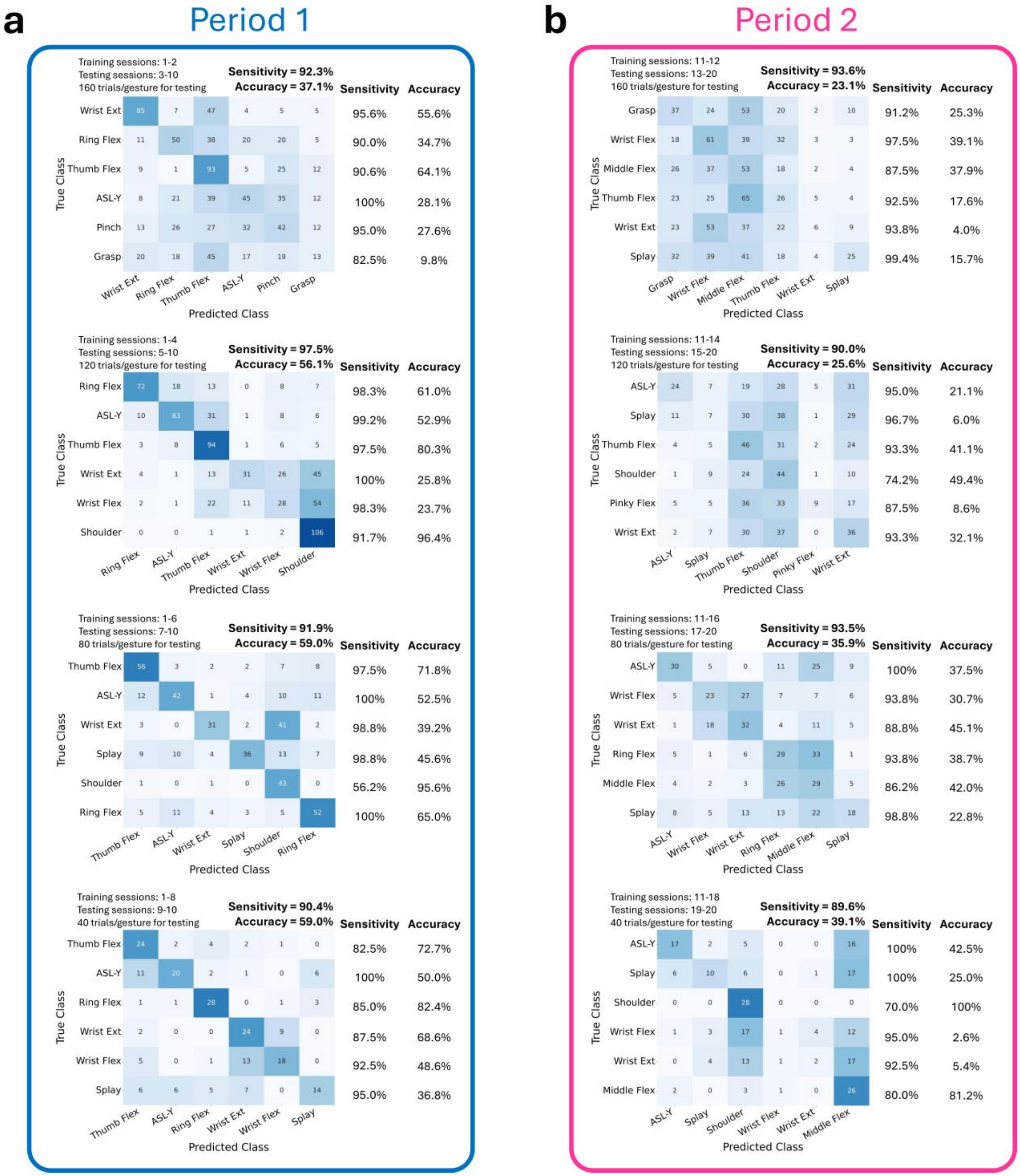

**Supplementary Figure 12 | Composite sensitivity, accuracy, and confusion matrices.** The composite sensitivities, accuracies, and confusion matrices are grouped by Period 1 and Period 2 (**a** and **b**, respectively). Each composite sensitivity and accuracy (bold, top right corner of each composite confusion matrix) represent the mean sensitivities and accuracies across the corresponding sets of testing sessions (top left corner of each composite confusion matrix). Each composite confusion matrix shows the element-wise sum of the individual confusion matrices for a particular set of testing sessions. The elements of the confusion matrices represent the number of predicted classifications, given the true classifications. For each composite confusion matrix, the color range is normalized to the respective number of trials per gesture for testing. The per-gesture composite sensitivity and composite accuracy are shown to the right of each composite confusion matrix.

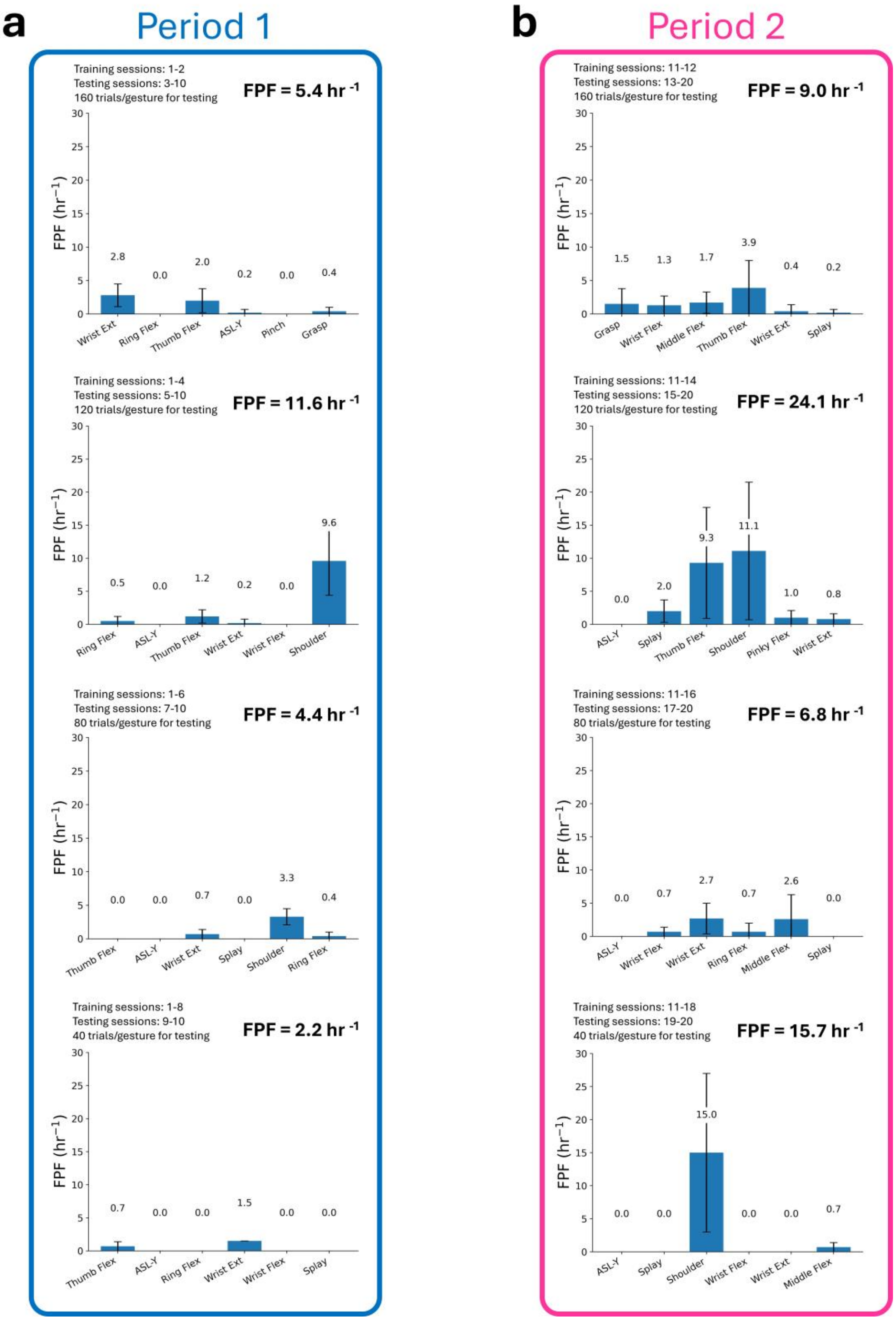


**Supplementary Figure 13 | Composite false positive frequencies (FPFs).** The composite FPF are grouped by Period 1 and Period 2 (**a** and **b**, respectively). Each composite FPF (bold, top right corner of each bar plot) represents the mean FPF across the corresponding sets of testing sessions (top left corner of each composite confusion matrix). For each set of testing sessions, the composite FPF is separated into the per-gesture FPF for that set of testing sessions.

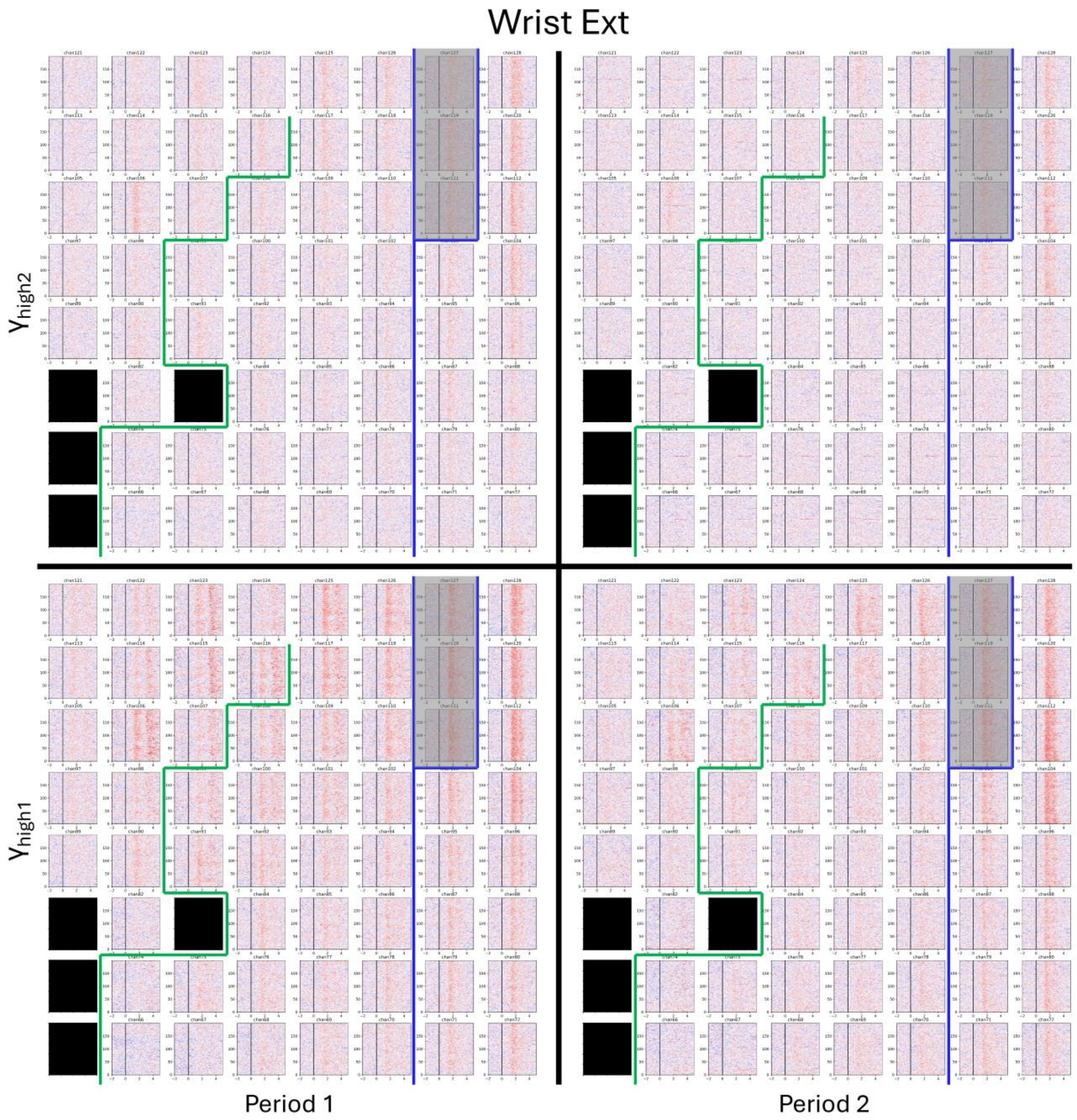


**Supplementary Figure 14 | Power trial rasters for Wrist Ext.** The aligned power trial rasters corresponding to modulation of the $\gamma_{high1}$ and $\gamma_{high2}$ frequency bands during attempted Wrist Ext movements are shown for Periods 1 and 2. Only signals from the grid covering the upper limb sensorimotor cortex are shown. The power trial rasters were computed and aligned as described in *Label assignment*. The power trial raster for each electrode is shown in each of the four panels. For any electrode, the vertical and horizontal axes represent the trial numbers and cue-aligned time segment (-2 to 5 s relative to visual cue onset). The vertical black line represents visual cue onset. The approximate central sulcus location is delineated by a thick blue line (CS) and widens at the top such that electrodes 111, 119, and 127 are over it. The pre-central sulcus is delineated by a thick green line (Pre-CS). The power trial rasters for electrodes 65, 73, 81, and 83 are not shown and marked in black because they were not used in analysis.

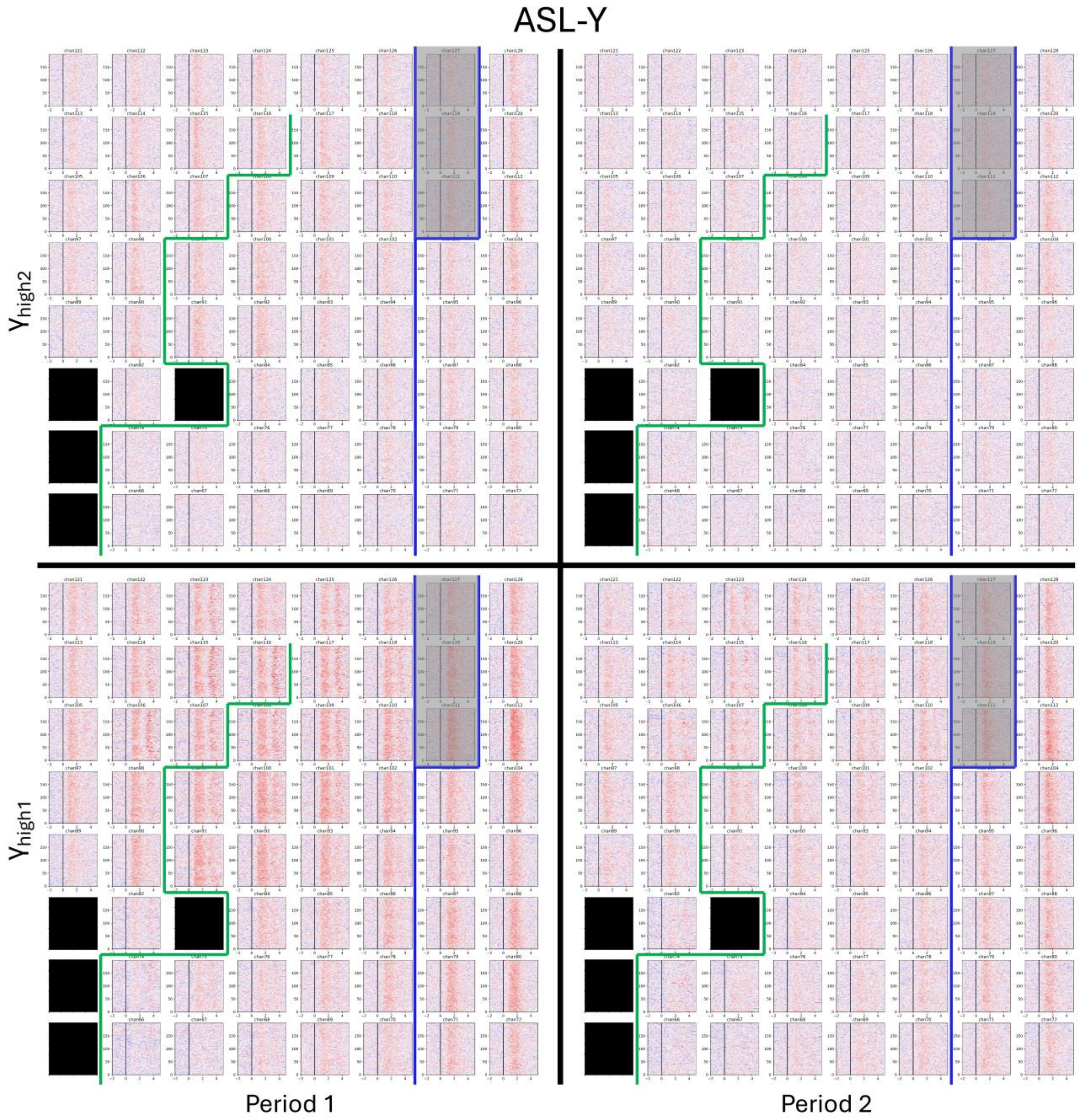


**Supplementary Figure 15 | Power trial rasters for ASL-Y.** The aligned power trial rasters corresponding to modulation of the $\gamma_{high1}$ and $\gamma_{high2}$ frequency bands during attempted ASL-Y movements are shown for Periods 1 and 2. Only signals from the grid covering the upper limb sensorimotor cortex are shown. The power trial rasters were computed and aligned as described in *Label assignment*. The power trial raster for each electrode is shown in each of the four panels. For any electrode, the vertical and horizontal axes represent the trial numbers and cue-aligned time segment (-2 to 5 s relative to visual cue onset). The vertical black line represents visual cue onset. The approximate central sulcus location is delineated by a thick blue line (CS) and widens at the top such that electrodes 111, 119, and 127 are over it. The pre-central sulcus is delineated by a thick green line (Pre-CS). The power trial rasters for electrodes 65, 73, 81, and 83 are not shown and marked in black because they were not used in analysis.

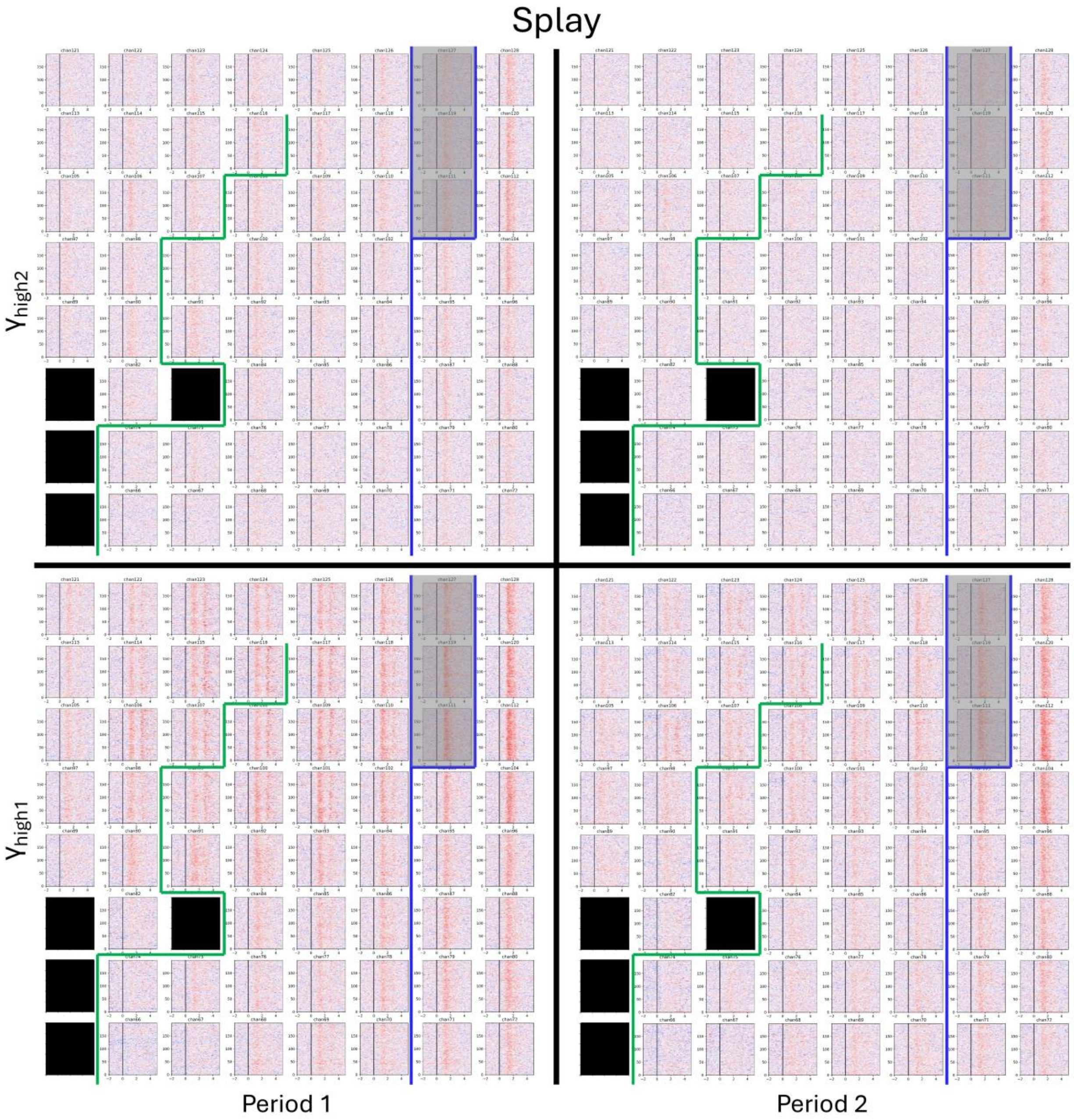


**Supplementary Figure 16 | Power trial rasters for Splay.** The aligned power trial rasters corresponding to modulation of the $\gamma_{high1}$ and $\gamma_{high2}$ frequency bands during attempted Splay movements are shown for Periods 1 and 2. Only signals from the grid covering the upper limb sensorimotor cortex are shown. The power trial rasters were computed and aligned as described in *Label assignment*. The power trial raster for each electrode is shown in each of the four panels. For any electrode, the vertical and horizontal axes represent the trial numbers and cue-aligned time segment (-2 to 5 s relative to visual cue onset). The vertical black line represents visual cue onset. The approximate central sulcus location is delineated by a thick blue line (CS) and widens at the top such that electrodes 111, 119, and 127 are over it. The pre-central sulcus is delineated by a thick green line (Pre-CS). The power trial rasters for electrodes 65, 73, 81, and 83 are not shown and marked in black because they were not used in analysis.

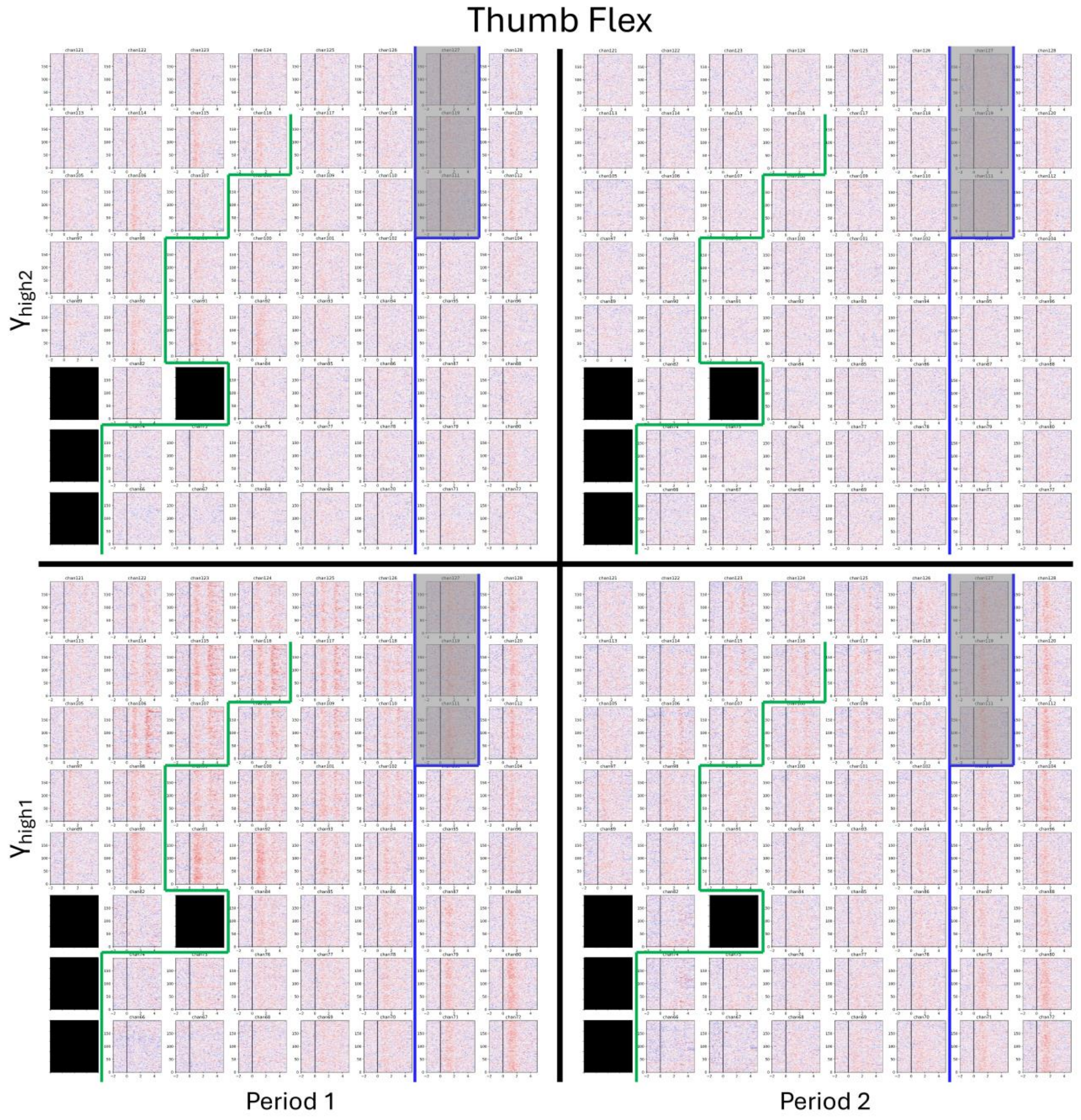


**Supplementary Figure 17 | Power trial rasters for Thumb Flex.** The aligned power trial rasters corresponding to modulation of the $\gamma_{high1}$ and $\gamma_{high2}$ frequency bands during attempted Thumb Flex movements are shown for Periods 1 and 2. Only signals from the grid covering the upper limb sensorimotor cortex are shown. The power trial rasters were computed and aligned as described in *Label assignment*. The power trial raster for each electrode is shown in each of the four panels. For any electrode, the vertical and horizontal axes represent the trial numbers and cue-aligned time segment (-2 to 5 s relative to visual cue onset). The vertical black line represents visual cue onset. The approximate central sulcus location is delineated by a thick blue line (CS) and widens at the top such that electrodes 111, 119, and 127 are over it. The pre-central sulcus is delineated by a thick green line (Pre-CS). The power trial rasters for electrodes 65, 73, 81, and 83 are not shown and marked in black because they were not used in analysis.

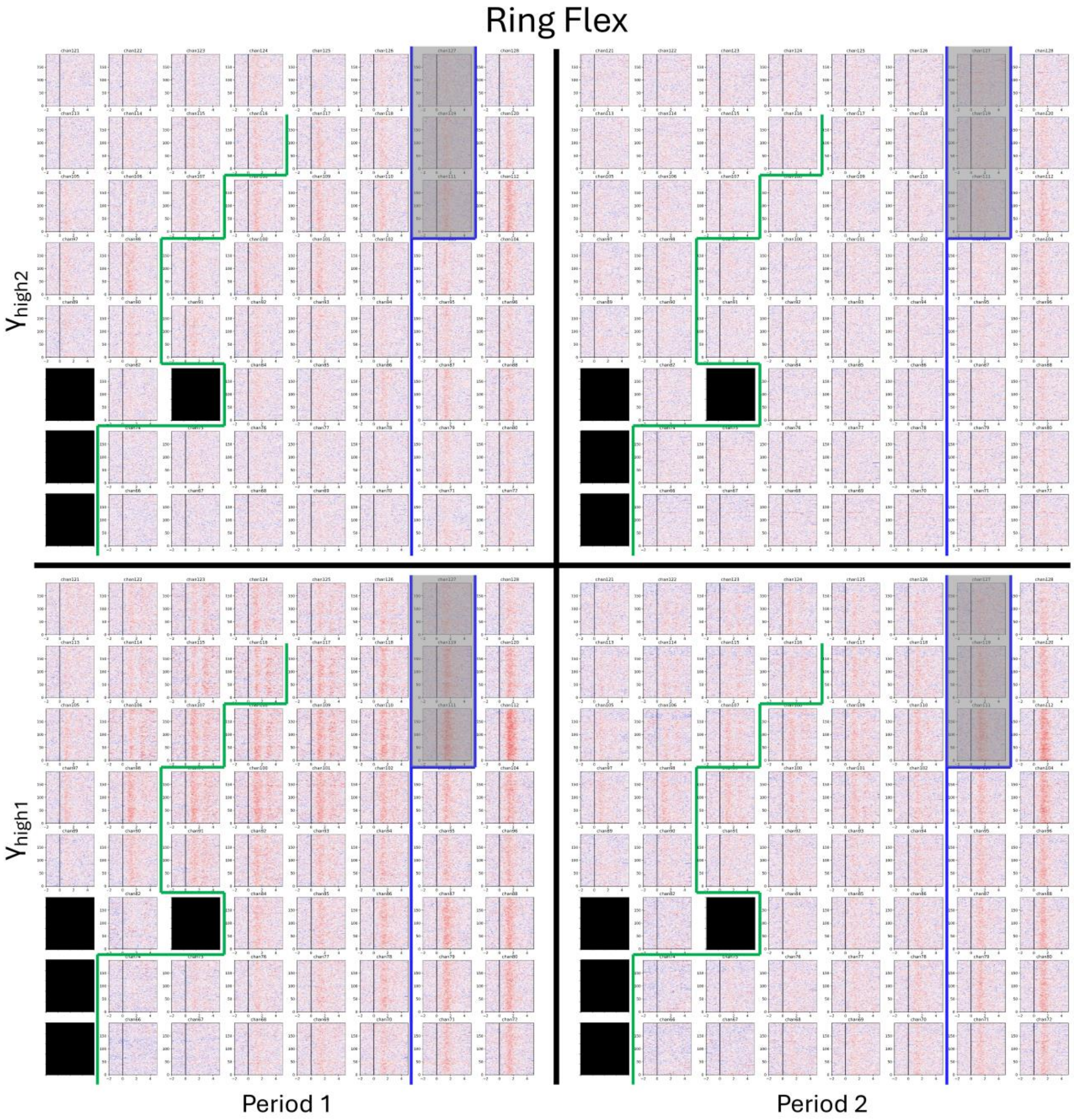


**Supplementary Figure 18 | Power trial rasters for Ring Flex.** The aligned power trial rasters corresponding to modulation of the $\gamma_{high1}$ and $\gamma_{high2}$ frequency bands during attempted Ring Flex movements are shown for Periods 1 and 2. Only signals from the grid covering the upper limb sensorimotor cortex are shown. The power trial rasters were computed and aligned as described in *Label assignment*. The power trial raster for each electrode is shown in each of the four panels. For any electrode, the vertical and horizontal axes represent the trial numbers and cue-aligned time segment (-2 to 5 s relative to visual cue onset). The vertical black line represents visual cue onset. The approximate central sulcus location is delineated by a thick blue line (CS) and widens at the top such that electrodes 111, 119, and 127 are over it. The pre-central sulcus is delineated by a thick green line (Pre-CS). The power trial rasters for electrodes 65, 73, 81, and 83 are not shown and marked in black because they were not used in analysis.

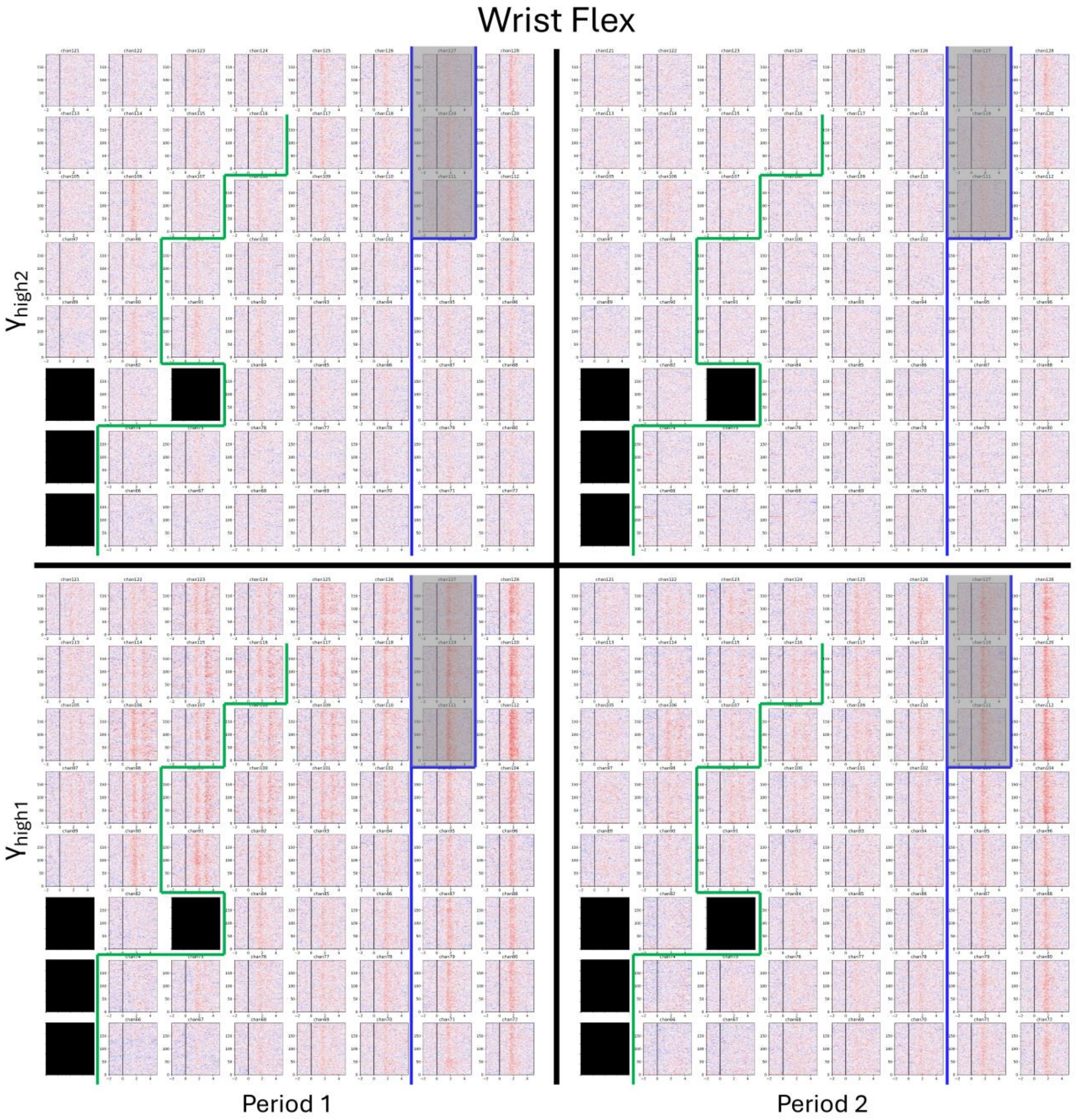


**Supplementary Figure 19 | Power trial rasters for Wrist Flex.** The aligned power trial rasters corresponding to modulation of the $\gamma_{high1}$ and $\gamma_{high2}$ frequency bands during attempted Wrist Flex movements are shown for Periods 1 and 2. Only signals from the grid covering the upper limb sensorimotor cortex are shown. The power trial rasters were computed and aligned as described in *Label assignment*. The power trial raster for each electrode is shown in each of the four panels. For any electrode, the vertical and horizontal axes represent the trial numbers and cue-aligned time segment (-2 to 5 s relative to visual cue onset). The vertical black line represents visual cue onset. The approximate central sulcus location is delineated by a thick blue line (CS) and widens at the top such that electrodes 111, 119, and 127 are over it. The pre-central sulcus is delineated by a thick green line (Pre-CS). The power trial rasters for electrodes 65, 73, 81, and 83 are not shown and marked in black because they were not used in analysis.

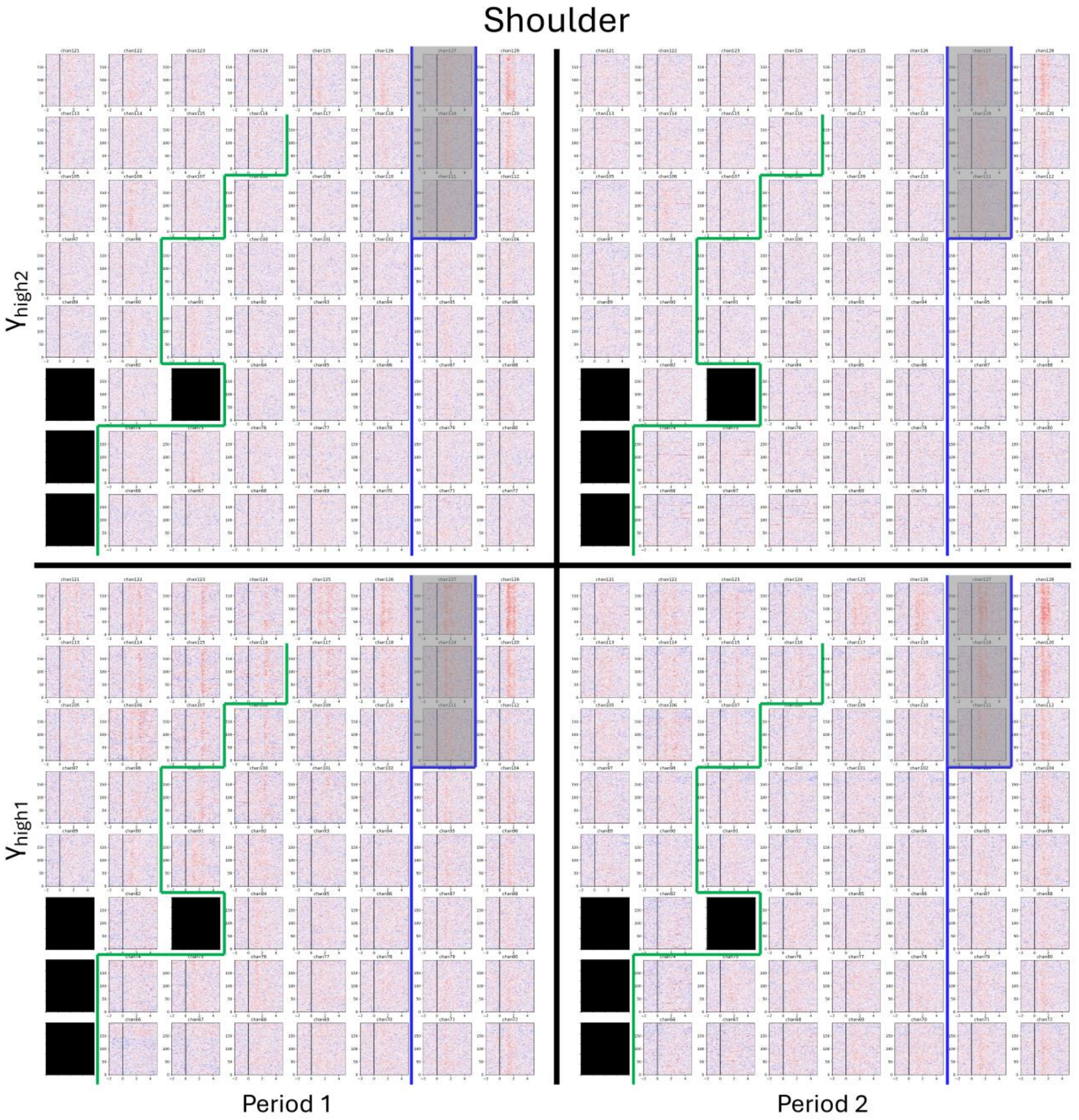


**Supplementary Figure 20 | Power trial rasters for Shoulder.** The aligned power trial rasters corresponding to modulation of the $\gamma_{high1}$ and $\gamma_{high2}$ frequency bands during attempted Shoulder movements are shown for Periods 1 and 2. Only signals from the grid covering the upper limb sensorimotor cortex are shown. The power trial rasters were computed and aligned as described in *Label assignment*. The power trial raster for each electrode is shown in each of the four panels. For any electrode, the vertical and horizontal axes represent the trial numbers and cue-aligned time segment (-2 to 5 s relative to visual cue onset). The vertical black line represents visual cue onset. The approximate central sulcus location is delineated by a thick blue line (CS) and widens at the top such that electrodes 111, 119, and 127 are over it. The pre-central sulcus is delineated by a thick green line (Pre-CS). The power trial rasters for electrodes 65, 73, 81, and 83 are not shown and marked in black because they were not used in analysis.

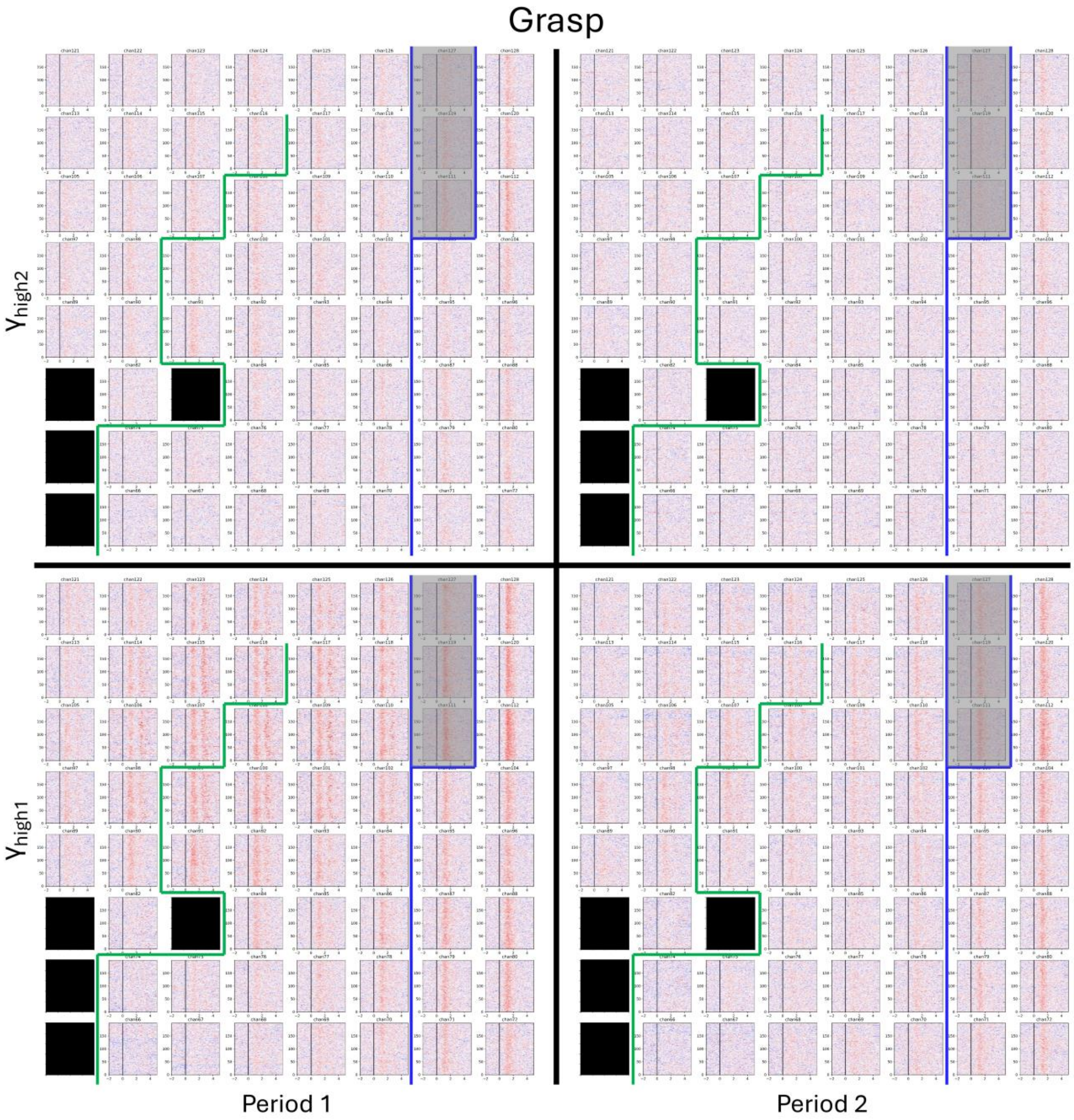


**Supplementary Figure 21 | Power trial rasters for Grasp.** The aligned power trial rasters corresponding to modulation of the $\gamma_{high1}$ and $\gamma_{high2}$ frequency bands during attempted Grasp movements are shown for Periods 1 and 2. Only signals from the grid covering the upper limb sensorimotor cortex are shown. The power trial rasters were computed and aligned as described in *Label assignment*. The power trial raster for each electrode is shown in each of the four panels. For any electrode, the vertical and horizontal axes represent the trial numbers and cue-aligned time segment (-2 to 5 s relative to visual cue onset). The vertical black line represents visual cue onset. The approximate central sulcus location is delineated by a thick blue line (CS) and widens at the top such that electrodes 111, 119, and 127 are over it. The pre-central sulcus is delineated by a thick green line (Pre-CS). The power trial rasters for electrodes 65, 73, 81, and 83 are not shown and marked in black because they were not used in analysis.

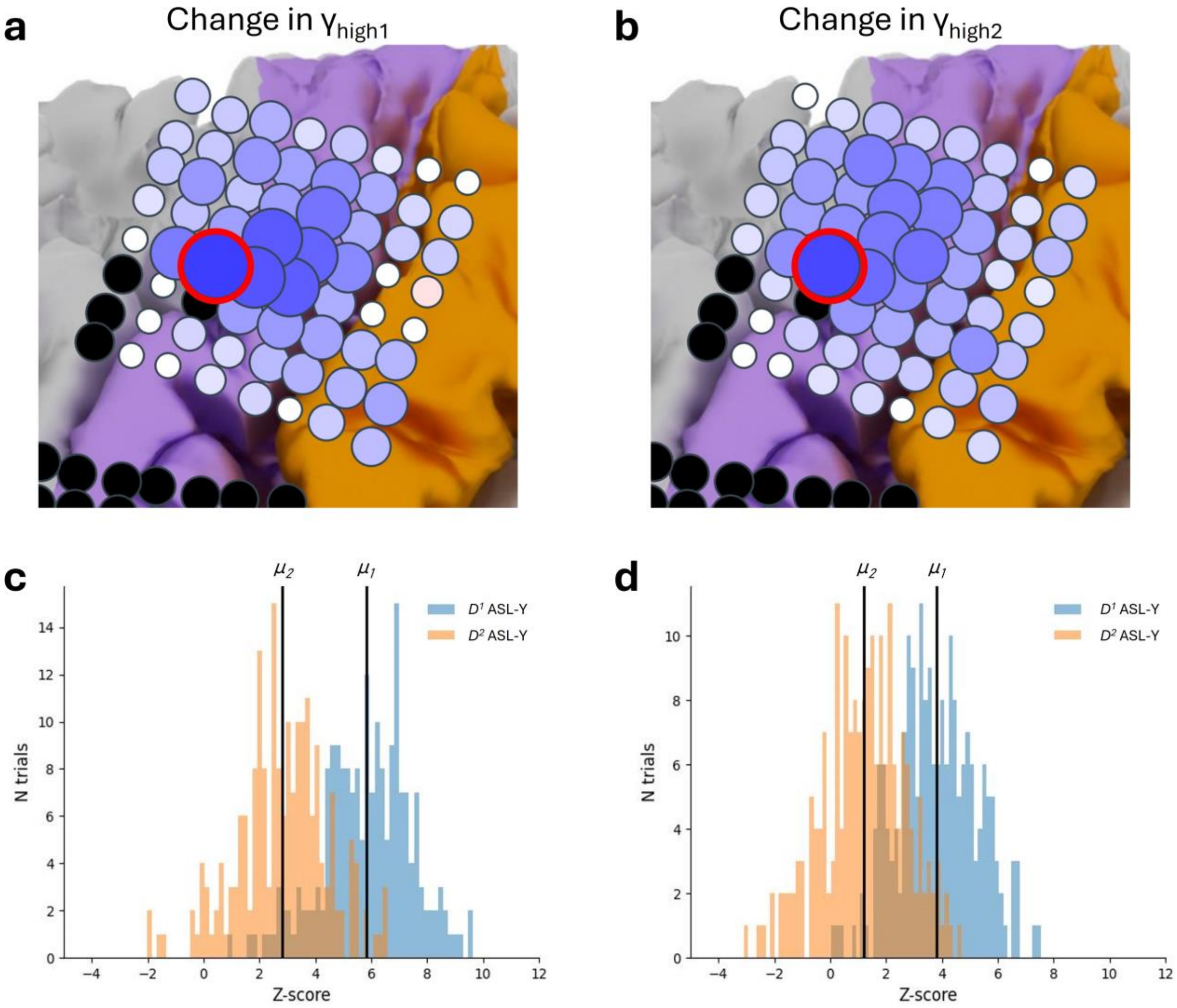


**Supplementary Figure 22 | Change in channel 91's high-gamma power during ASL-Y.** For ASL-Y, the changes in $\gamma_{high1}$ (**a**) and $\gamma_{high2}$ (**b**) modulation from Period 1 and Period 2 across all electrodes are shown and overlayed on a schematic of the left hemisphere (replicated from Fig. 4 in the main text). Channel 91 is circled in red. The differences in channel 91's modulation of $\gamma_{high1}$ and $\gamma_{high2}$ power between the Periods 1 and 2 are shown in the histograms of (**c**) and (**d**), respectively. Color coding of the cortex indicates anatomical landmarks: magenta for pre-central gyrus and orange for post-central gyrus. (**c**, **d**) For each powerband the $D^1$ distribution is composed of each trial's mean activity between the corresponding time points of the 70% threshold crossing of Period 1 (Supplementary Fig 11h). Similarly, the $D^2$ distribution is computed but using the corresponding time points of Period 2. Since spectral activity was standardized to baseline, the powerband activity has standard units. For each powerband, $\mu_1$ and $\mu_2$ are the means of the distributions. For $\gamma_{high1}$, $\mu_1 = 5.8$ and $\mu_2 = 2.8$. For $\gamma_{high2}$, $\mu_1 = 3.8$ and $\mu_2 = 1.2$. For the $\gamma_{high1}$ and $\gamma_{high2}$ powerband the $D^2$ distribution is significantly lower than the $D^1$ distribution (*P* = 2.5 x $10^{-57}$ and *P* = 1.1 x $10^{-52}$, Welch's t-test for both comparisons).

### Supplementary Tables

| Movement | Visible | Instruction | Observation | Correctly executed? |
|---|---|---|---|---|
| Grasp | Yes | Flex all your fingers of your right hand together in a fist. | Flexion of the right middle and ring fingers. | Partial |
| ASL-Y | Yes | Flex your right index, middle, and ring fingers while extending the thumb and pinky fingers. | Flexion of the middle and ring fingers. Slight extension of the pinky finger. | Partial |
| Splay | Yes | Spread all five fingers of your right hand radially outward such that the tip of the thumb and pinky are as far from each other as possible. | Slight radial spreading of all right fingers. Slight extension of middle, ring, and pinky fingers as well as palm. At maximum spread, angle between palm/fingers and floor is roughly 45 degrees. Index finger and thumb do not extend. | Partial |
| Pinch | Yes* | Bring your tips of the right thumb and index finger together while keeping the other fingers still. | Flexion of the right middle and ring fingers. *No visible movement of the right thumb. | None |
| Wrist Ext | Yes | Extend your right hand as much as possible. | Extension of the right hand as well as middle, ring, and pinky fingers of the right hand. At maximum extension angle between right hand palm and floor is roughly 30 degrees. | Partial |
| Wrist Flex | Yes | Flex your right hand inward as far as possible. | Flexion of the middle and ring fingers. Only slight flexion at the wrist. | Partial |
| Thumb Flex | Yes* | Flex your right thumb toward the palm of your hand. | Slight movement of entire right hand in lateral direction. *No visible movement of the thumb. | None |
| Index Flex | Yes* | Flex your right index finger inward toward the palm while keeping the other fingers still. | Flexion of the middle and ring fingers. *No visible movement of the index finger. | None |
| Middle Flex | Yes | Flex your right middle finger inward toward your palm while keeping the other fingers still. | Flexion of the right middle and ring fingers. | Partial |
| Ring Flex | Yes | Flex your right ring finger inward toward your palm while keeping the other fingers still. | Flexion of the right middle and ring fingers. | Partial |
| Pinky Flex | Yes* | Flex your right pinky finger inward toward the palm while keeping the other fingers still. | Flexion of the right middle and ring fingers. *No visible movement of the pinky finger. | None |
| Heel Raise | Yes | Raise the heel of your right foot while keeping the balls of the foot planted. | Right heel raised with high range of motion. Balls of the foot planted firmly. | Complete |
| Shoulder | Yes | Shrug your right shoulder upward toward your right ear. | Shrugged right shoulder upward. Half range of motion. | Partial |
| Grimace | Yes | Extend the corners of your mouth lateral and part lips to show closed bite. | Corners of the mouth extend laterally and downward. Lips part to show teeth in a closed bite. | Complete |
| Tongue | No | Touch your tongue to the back of your front teeth without opening your mouth. | None. | Complete |
| Eyebrows | Yes | Raise your eyebrows. | Raised eyebrows. | Complete |

**Supplementary Table 1 | Descriptions of attempted gestures.** For each of the 16 visual gesture stimuli displayed on the monitor, the instructions on how to perform the movement, whether the movement was physically observable, and the description of the observable movement are detailed above. Ext = Extension; Flex = Flexion. * represents a visible movement, though not necessarily the correct one.

| | Model | Training sessions | Testing sessions | Sensitivity (%) mean ± 95% CI | Accuracy (%) mean ± 95% CI | FPF (hr $^{-1}$) mean ± 95% CI | Latency (s) mean ± 95% CI |
|---|---|---|---|---|---|---|---|
| $D^1$ training sessions, $D^1$ testing sessions | Model 1 | 1-2 | 3-10 | 92.3 ± [87.4, 97.2] | 37.1 ± [33.4, 40.9] | 6.8 ± [4.0, 9.5] | 1.16 ± [1.15, 1.18] |
| | Model 2 | 1-4 | 5-10 | 97.5 ± [96.1, 98.9] | 56.2 ± [50.8, 61.6] | 14.6 ± [6.9, 22.3] | 1.23 ± [1.21, 1.25] |
| | Model 3 | 1-6 | 7-10 | 91.9 ± [85.5, 98.2] | 58.9 ± [50.9, 66.8] | 5.6 ± [3.2, 8.0] | 1.12 ± [1.09, 1.14] |
| | Model 4 | 1-8 | 9-10 | 92.1 ± [n/a, n/a] | 67.8 ± [n/a, n/a] | 2.8 ± [n/a, n/a] | 1.29 ± [1.22, 1.3] |
| $D^2$ training sessions, $D^2$ testing sessions | Model 5 | 11-12 | 13-20 | 93.7 ± [90.2, 97.1] | 23.2 ± [18.6, 27.7] | 11.5 ± [4.0, 14.2] | 1.19 ± [1.17, 1.21] |
| | Model 6 | 11-14 | 15-20 | 90.0 ± [86.9, 93.1] | 25.6 ± [24.0, 27.2] | 30.6 ± [6.5, 41.9] | 1.15 ± [1.12, 1.17] |
| | Model 7 | 11-16 | 17-20 | 93.5 ± [88.8, 98.3] | 35.7 ± [26.2, 45.3] | 8.6 ± [1.8, 11.8] | 1.26 ± [1.23, 1.29] |
| | Model 8 | 11-18 | 19-20 | 89.6 ± [n/a, n/a] | 39.1 ± [n/a, n/a] | 19.9 ± [n/a, n/a] | 1.20 ± [1.16, 1.24] |

**Supplementary Table 2 | Offline performance per model.** For each set of training sessions, the resulting model was tested on the corresponding set of testing sessions. The performance metrics (sensitivity, accuracy, false positive frequency (FPF), and latency) of each model shown in Fig. 2a, b, d, e, g, h, j, and k of the main text are represented by the mean and 95% confidence interval (CI).

#### Sensitivity

| | Means (%) 95% CI |
|---|---|
| $D^1$ train $D^1$ test | 93.6 [91.3, 95.9] |
| $D^2$ train $D^2$ test | 92.1 [90.4, 93.8] |
| Is $P < 0.05$ | No |

#### Accuracy

| | Means (%) 95% CI |
|---|---|
| $D^1$ train $D^1$ test | 49.3 [44.1, 54.6] |
| $D^2$ train $D^2$ test | 28.0 [24.5, 31.5] |
| Is $P < 0.05$ | $2.1 \times 10^{-6}$ |

#### False positive frequency

| | Means (hr $^{-1}$) 95% CI |
|---|---|
| $D^1$ train $D^1$ test | 8.5 [5.6, 11.3] |
| $D^2$ train $D^2$ test | 17.5 [8.6, 25.4] |
| Is $P < 0.05$ | $3.9 \times 10^{-2}$ |

#### Latency

| | Means (s) 95% CI |
|---|---|
| $D^1$ train $D^1$ test | 1.18 [1.15, 1.21] |
| $D^2$ train $D^2$ test | 1.19 [1.16, 1.23] |
| Is $P < 0.05$ | No |

**Supplementary Table 3 | Summary of offline performance.** The mean performance metrics across all models trained and tested using data exclusively from $D^1$ and using data exclusively from $D^2$ that are shown in Fig. 2c, f, I, and l in the main text. The significant $P$-values are shown for the comparison of each performance metric (Sensitivity, Accuracy, FPF, and Latency are shown in **a**-**d** respectively) between models trained and tested using data exclusively from $D^1$ and using data exclusively from $D^2$. For $P$-values that were non-significant, “No” is shown.

**a** Sensitivity

| Means (%) 95% CI | Wrist Ext | ASL-Y | Splay | Thumb Flex | Ring Flex | Wrist Flex | Shoulder | Grasp | Pinch | Pinky Flex | Middle Flex |
|---|---|---|---|---|---|---|---|---|---|---|---|
| $D^1$ train $D^1$ test | 96.3 [93.7, 99.0] | 99.8 [99.2, 100] | 97.5 [94.6, 100] | 92.1 [87.1, 97.1] | 93.9 [90.0, 97.9] | 96.4 [92.9, 99.9] | 75.6 [59.0, 92.1] | 85.0 [73.6, 96.4] | 95.0 [89.5, 100] | n/a | n/a |
| $D^2$ train $D^2$ test | 92.5 [89.6, 95.4] | 97.5 [94.6, 100] | 98.5 [97.2, 99.8] | 92.9 [89.0, 96.7] | 93.8 [86.1, 100] | 96.1 [93.5, 98.6] | 73.1 [61.5, 84.7] | 91.2 [84.6, 97.9] | n/a | 84.2 [75.1, 93.2] | 86.1 [81.7, 90.5] |
| Is $P < 0.05$? | $4.0 \times 10^{-2}$ | No | No | No | No | No | No | No | n/a | n/a | n/a |

**b** Accuracy

| Means (%) 95% CI | Wrist Ext | ASL-Y | Splay | Thumb Flex | Ring Flex | Wrist Flex | Shoulder | Grasp | Pinch | Pinky Flex | Middle Flex |
|---|---|---|---|---|---|---|---|---|---|---|---|
| $D^1$ train $D^1$ test | 46.4 [38.1, 54.6] | 42.6 [34.2, 51.1] | 42.7 [37.8, 47.5] | 71.8 [66.1, 77.5] | 53.3 [43.4, 63.3] | 31.4 [16.4, 46.4] | 95.2 [89.7, 100] | 9.7 [4.1, 15.3] | 28.1 [17.0, 39.2] | n/a | n/a |
| $D^2$ train $D^2$ test | 20.8 [12.0, 29.6] | 30.1 [18.5, 41.8] | 15.1 [10.6, 19.6] | 27.4 [17.2, 37.6] | 38.6 [0.6, 76.5] | 31.5 [21.6, 41.5] | 62.4 [41.8, 83.0] | 25.5 [16.3, 34.8] | n/a | 8.1 [0, 16.8] | 45.7 [35.1, 56.2 |
| Is $P < 0.05$? | $3.8 \times 10^{-4}$ | No | $2.6 \times 10^{-4}$ | $3.4 \times 10^{-6}$ | No | $3.4 \times 10^{-6}$ | $2.7 \times 10^{-2}$ | $4.6 \times 10^{-3}$ | n/a | n/a | n/a |

**c** False positive frequency

| Means (hr$^{-1}$) 95% CI | Wrist Ext | ASL-Y | Splay | Thumb Flex | Ring Flex | Wrist Flex | Shoulder | Grasp | Pinch | Pinky Flex | Middle Flex |
|---|---|---|---|---|---|---|---|---|---|---|---|
| $D^1$ train $D^1$ test | 2.1 [1.1, 3.0] | 0.1 [0, 0.3] | 0 [0, 0] | 1.4 [0.4, 2.3] | 0.2 [0, 0.5] | 0 [0, 0] | 8.5 [3.0, 14.0] | 0.5 [0, 1.2] | 0 [0, 0] | n/a | n/a |
| $D^2$ train $D^2$ test | 1.2 [0.3, 2.2] | 0 [0, 0] | 0.9 [0.1, 1.7] | 7.9 [2.7, 13.1] | 0.9 [0, 3.9] | 1.1 [0.1, 2.0] | 15.2 [2.9, 27.5] | 1.9 [0, 4.5] | n/a | 1.9 [0, 4.2] | 2.3 [0.5, 4.2] |
| Is $P < 0.05$? | No | No | No | $2.8 \times 10^{-3}$ | No | No | No | No | n/a | n/a | n/a |

**d** Latency

| Means (s) 95% CI | Wrist Ext | ASL-Y | Splay | Thumb Flex | Ring Flex | Wrist Flex | Shoulder | Grasp | Pinch | Pinky Flex | Middle Flex |
|---|---|---|---|---|---|---|---|---|---|---|---|
| $D^1$ train $D^1$ test | 1.41 [1.36, 1.47] | 1.05 [1.02, 1.07] | 0.91 [0.83, 1.00] | 1.12 [1.08, 1.15] | 1.16 [1.12, 1.20] | 1.46 [1.34, 1.58] | 1.40 [1.36, 1.45] | 1.06 [1.03, 1.10] | 1.06 [1.0, 1.11 | n/a | n/a |
| $D^2$ train $D^2$ test | 1.36 [1.31, 1.42] | 1.04 [0.98, 1.10] | 1.00 [0.96, 1.05] | 1.16 [1.11, 1.21] | 1.32 [1.25, 1.39] | 1.49 [1.42, 1.55] | 1.18 [1.13, 1.24] | 1.03 [0.97, 1.09] | n/a | 1.26 [1.19, 1.34] | 1.14 [1.11, 1.17] |
| Is $P < 0.05$? | No | No | No | No | $3.5 \times 10^{-3}$ | No | No | No | n/a | n/a | n/a |

**Supplementary Table 4 | Performance metrics per gesture.** The mean performance metrics for each gesture across all models trained and tested using data exclusively from $D^1$ and using data exclusively from $D^2$ that are shown in Fig. 3 of the main text. The mean sensitivity (**a**), accuracy (**b**), false positive frequency (FPF) (**c**), and latency metrics (**d**) are shown along with a 95% confidence interval (CI). “n/a” represents gestures that were never used for models trained and tested on data from $D^1$ or from $D^2$. The significant *P*-values are shown for the comparison of each performance metric for each gesture between models trained and tested using data exclusively from $D^1$ and using data exclusively from $D^2$. For *P*-values that were non-significant or for comparisons that could not be made due to a gesture not used in either Period 1 or Period 2, “No” or “n/a” is shown.

### References

1. Cedarbaum, J. M. *et al.* The ALSFRS-R: a revised ALS functional rating scale that incorporates assessments of respiratory function. *Journal of the Neurological Sciences* **169**, 13–21 (1999).

2. Natraj, N. *et al.* Sampling representational plasticity of simple imagined movements across days enables long-term neuroprosthetic control. *Cell* **188**, 1208-1225.e32 (2025).

3. Williams, A. H. *et al.* Discovering Precise Temporal Patterns in Large-Scale Neural Recordings through Robust and Interpretable Time Warping. *Neuron* **105**, 246-259.e8 (2020).
